# Digital Health Interventions in Palliative Care: A Systematic Meta-Review and Evidence Synthesis

**DOI:** 10.1101/2020.09.16.20195834

**Authors:** Anne M. Finucane, Hannah O’Donnell, Jean Lugton, Connie Swenson, Claudia Pagliari

## Abstract

Digital health interventions (DHIs) have the potential to improve the accessibility and effectiveness of palliative care but heterogeneity amongst existing systematic reviews presents a challenge for evidence synthesis. This rigorous meta-review applied a structured search of 10 databases from 2006 to 2020, revealing 21 relevant systematic reviews, encompassing 332 unique publications. Most reviews were moderate quality. Interventions delivered via videoconferencing (17%), electronic healthcare records (16%) and phone (13%) were most frequently described. DHIs were typically used in palliative care for education (20%), symptom management (15%), decision-making support (13%), information provision or management (13%), and communication (9%). Positive impacts were reported on education, decision-making, information-sharing, communication, and costs. Impacts on symptom management were either positive or showed no harmful effects. However often DHIs were described but not evaluated. Responsive pragmatic research designs are now needed to guide further evaluation, implementation and to inform future service innovation.

---

The diagnosis of a life-limiting illness, along with its management during periods of wellness, illness, remission, decline and end-of-life can be stressful for patients, caregivers, and healthcare professionals. Palliative care offers a holistic set of approaches for ameliorating the physical, psychological, social and spiritual burdens patients and their families face.^1,2^ Improving access to, and increasing the quality of palliative care delivered is a healthcare priority in many countries.^3,4^ Digital health interventions (DHIs) could have an essential role to play in achieving these aims.

Digital health, or eHealth, is a broad term used to refer to the application of information and communication technologies (ICTs) and networks for the management, delivery and optimisation of patient care and health services, and for supporting patients themselves. It encompasses a range of related concepts such as telemedicine and telehealth, mobile (m)Health, health informatics, and wearable devices.^5,6^ The adoption of digital health technologies is rapidly changing how healthcare is provided. Electronic health records (EHRs) and decision support tools are part of routine healthcare practice in many countries. Mobile phones, apps, wearables and social media are in widespread use, and innovations such as augmented reality, virtual assistants and artificial intelligence are finding new uses in clinical management and patient self-care. These approaches are reshaping healthcare as they become more affordable and widespread.^7^

Palliative care is one area where these technologies are increasingly being deployed.^8^ Research to establish the feasibility of using videoconferencing in palliative care was first reported nearly 20 years ago.^9^ In healthcare organisations, pathways and preferences for palliative care are being steadily integrated into EHRs.^10^ In parallel, mobile applications and online social networks for supporting patients’ physical, cognitive and emotional needs are becoming popular, both supplied by healthcare providers^11^ and driven by patients and carers themselves.^12^ More recently, predictive analytics and artificial intelligence are being used to adapt clinical interventions to stages of terminal illness.^13^

Reflecting this activity, there has been a significant rise in the number of systematic reviews focused on DHIs and palliative care over the past 15 years.^14-18^ Despite their general support for these approaches, the clinical scope and quality of existing reviews varies widely, making it difficult to evaluate their implications for the field as a whole. Given the growing demand for palliative care services worldwide^19^ and the increasing penetration of DHIs in healthcare, the time is right for a comprehensive synthesis and appraisal of this evidence base. We employed the meta-review method to capture, appraise and synthesise the evidence represented in the systematic review literature on DHIs in palliative care. Our objectives were:

1. To identify the range of palliative DHIs described in existing systematic reviews.
2. To describe the quality of existing systematic review evidence.
3. To synthesize evidence on the role and the effects of DHIs in palliative care.
4. To identify evidence gaps and make recommendations for future research.

## RESULTS

### Search results

The database searches returned a total of 5092 titles and abstracts, of which 55 potentially relevant papers were subjected to full-text review and 21 were eligible for inclusion **(Figure 1)**. The main reason for excluding articles at full text review was that they were not focused on palliative care (13 studies) or DHIs (7 studies); not systematic reviews (4 studies); reviews of apps as opposed to research (3 studies); did not report on effects of DHIs or provide detail on included reviews (4 studies) or other reasons (3 studies). During the search process we identified one meta-review of telemedicine in palliative care published in 2016.^20^ This meta-review identified a total of 6 systematic reviews published between 2007 and 2012, all of which were included amongst the 21 eligible reviews in this meta-review.

**Figure 1:**
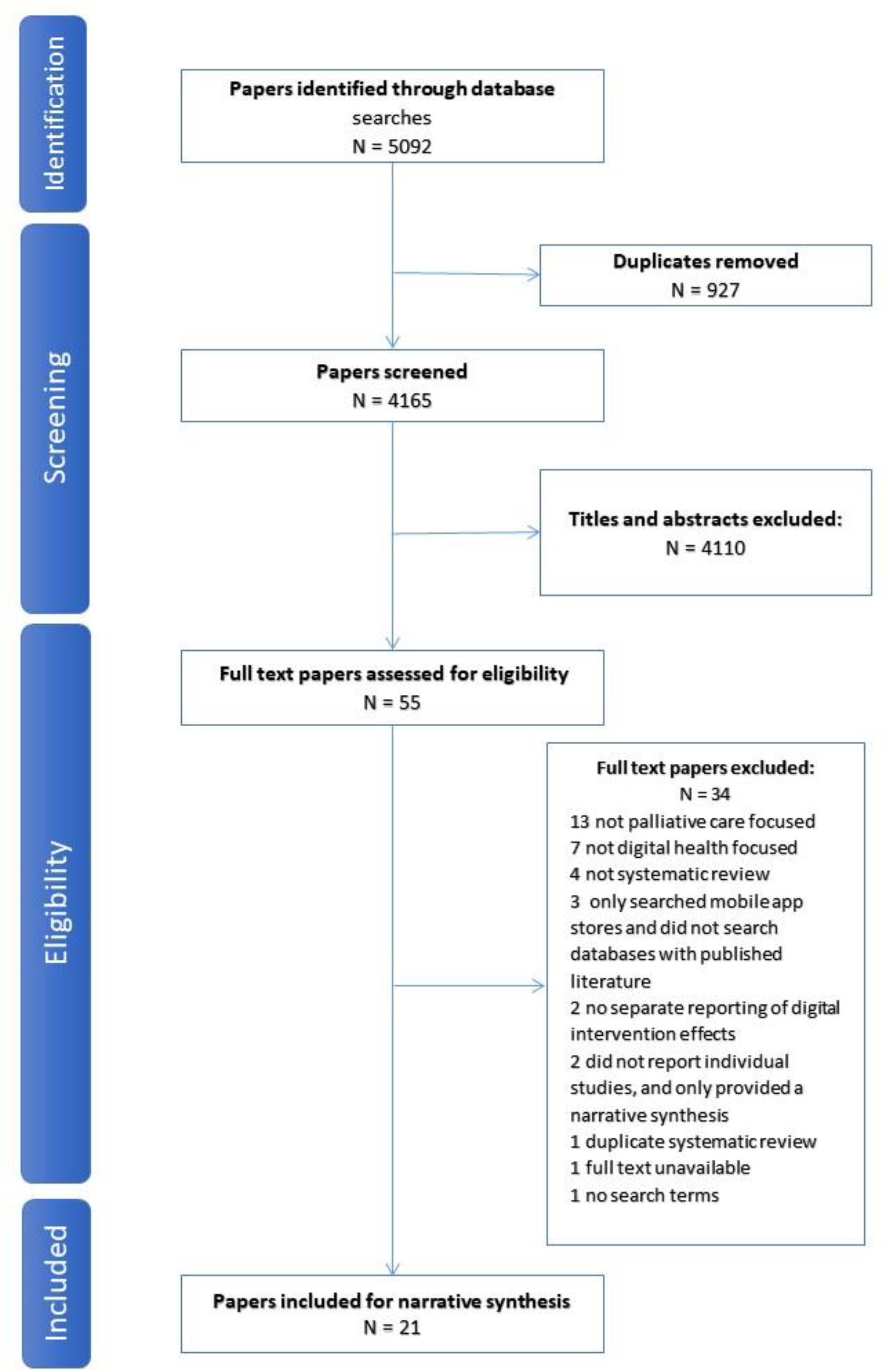
Prisma diagram.

### Description of the included studies

#### Characteristics of the included reviews

Characteristics of the 21 included reviews are shown in **Table 1 (See Appendix)**. Most included a range of study populations - patients, family members, caregivers, and health professionals. Three reviews focused on cancer,^21-23^ others did not limit their inclusion criteria a specific disease. The reviews were carried out by research teams based in the following countries: USA (9),^17,24-31^ UK (6),^16,23,32-35^ Australia (2),^14,36^ Canada (1)^22^ Chile (1),^15^ Denmark (1)^37^ and Brazil (1)^38^.

**Table 1:**
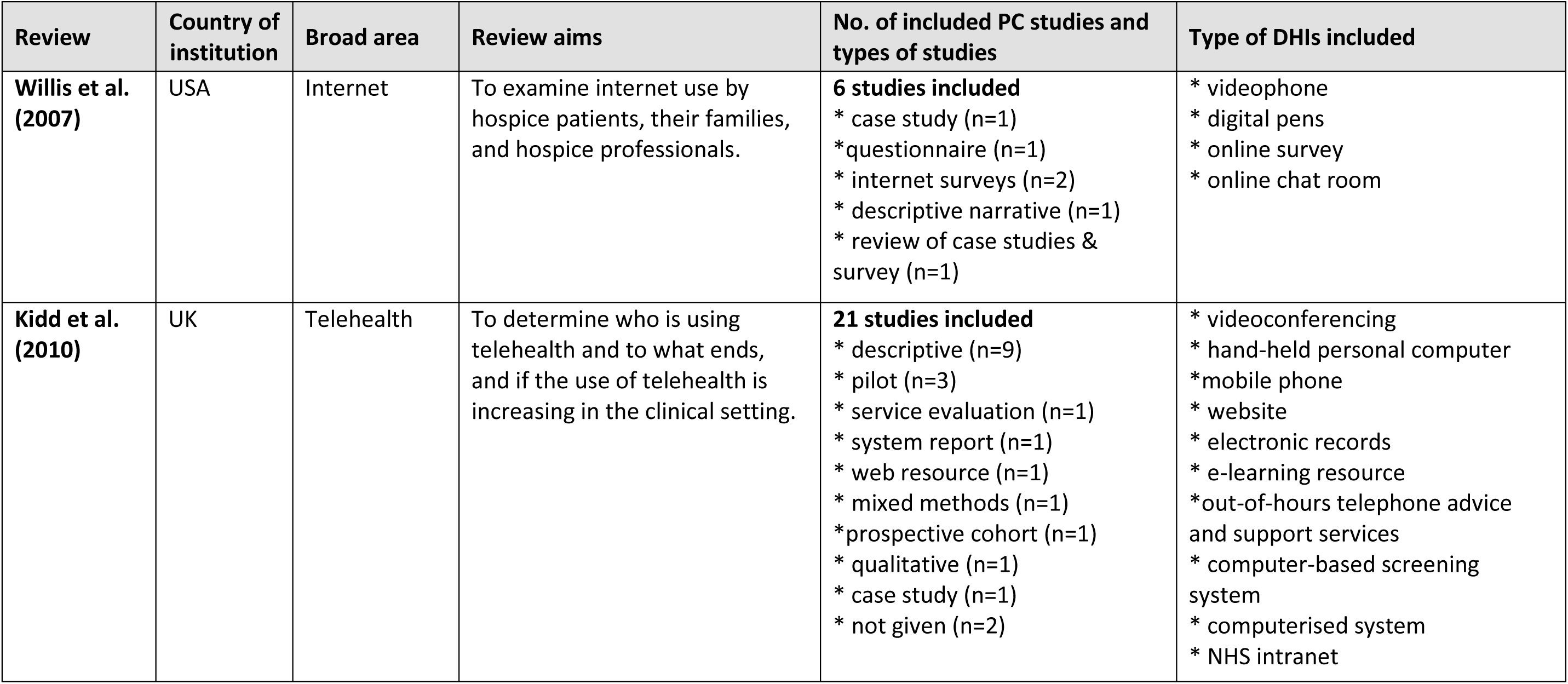
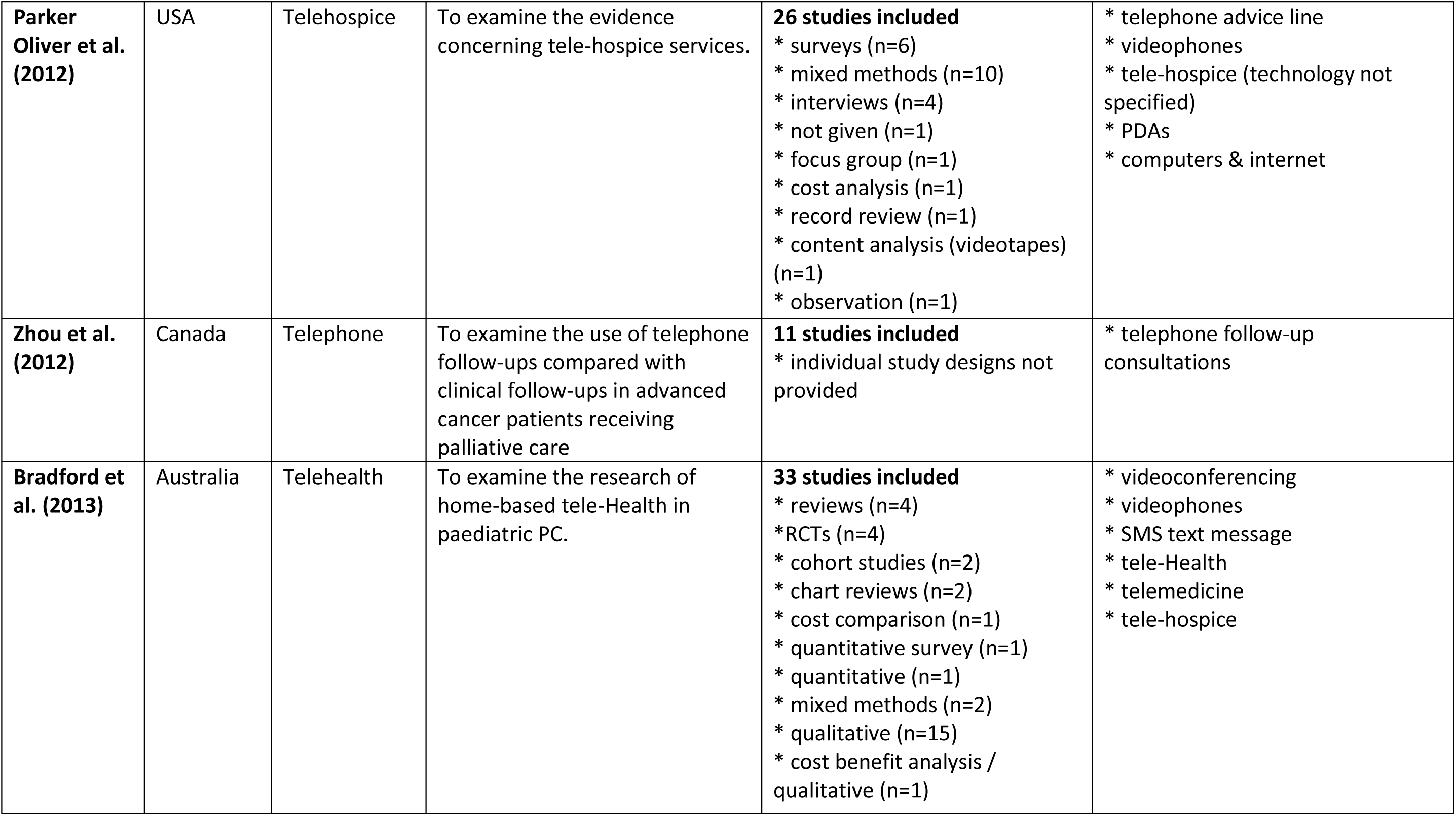
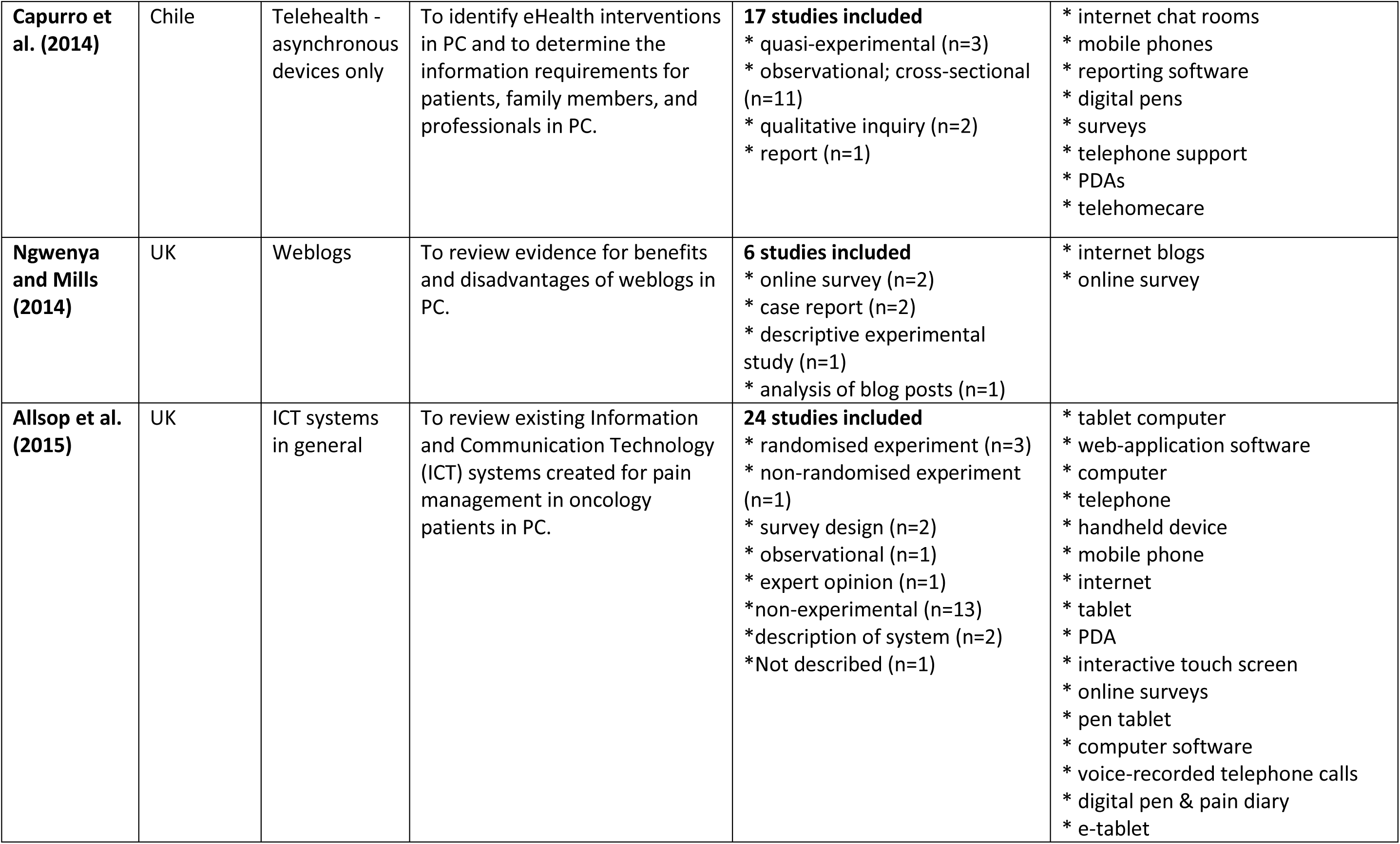
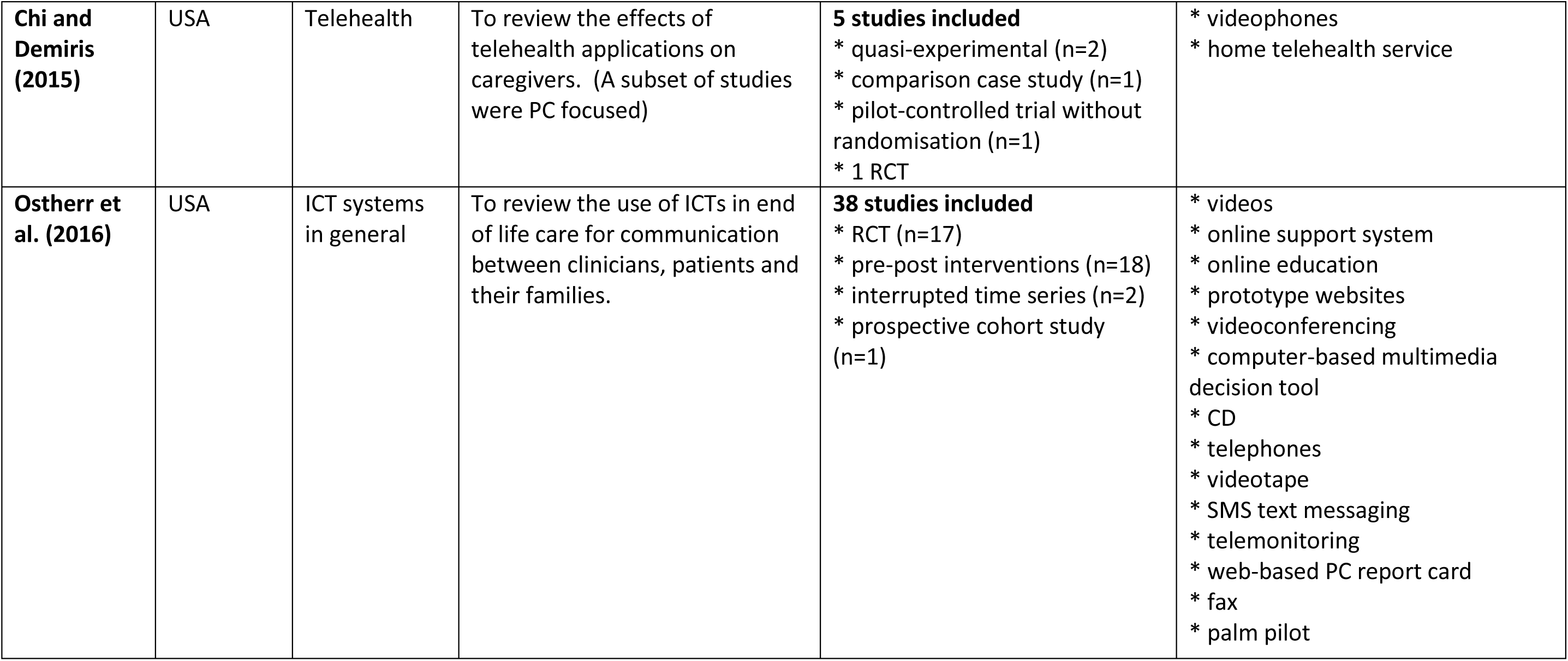
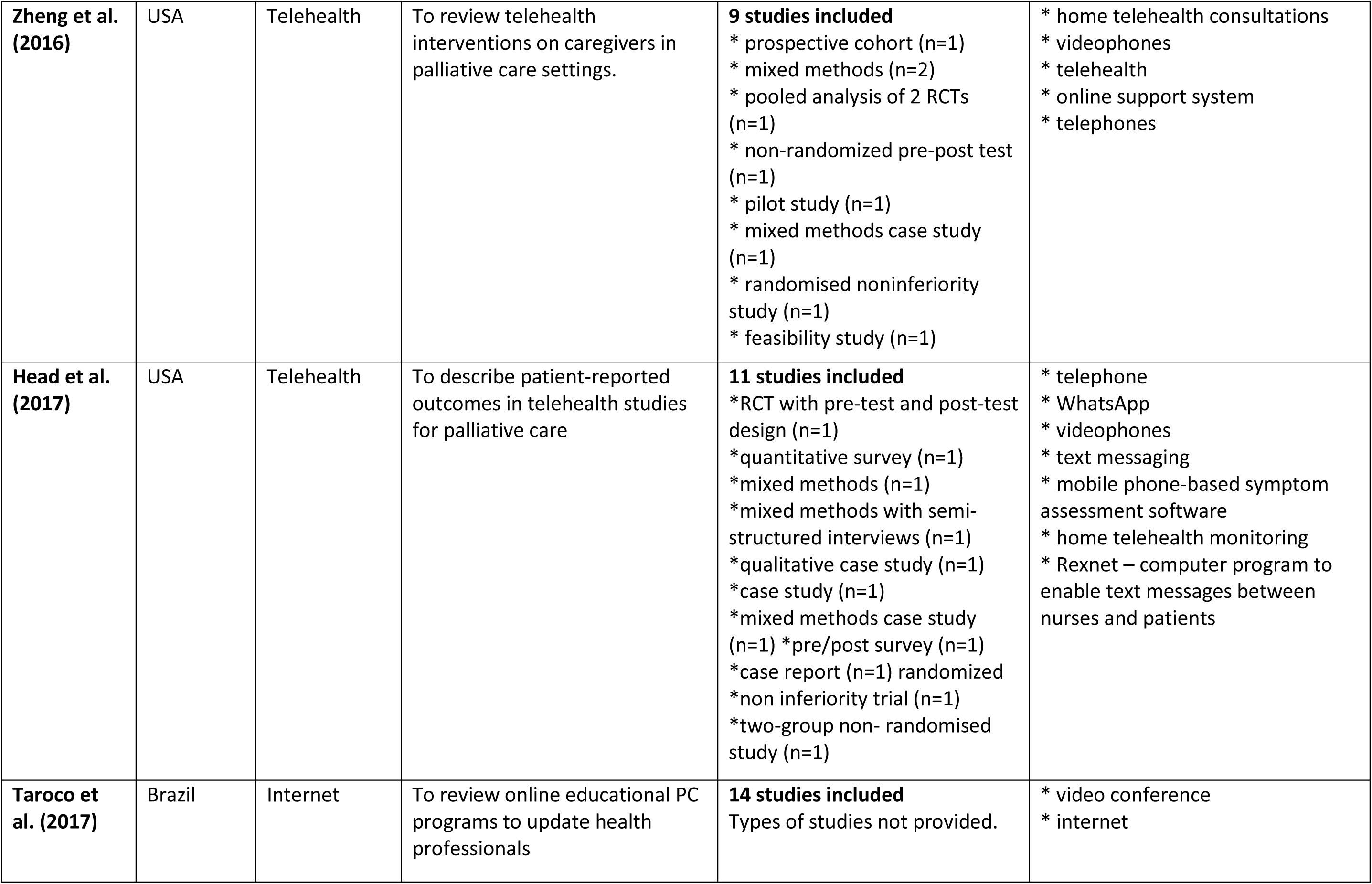
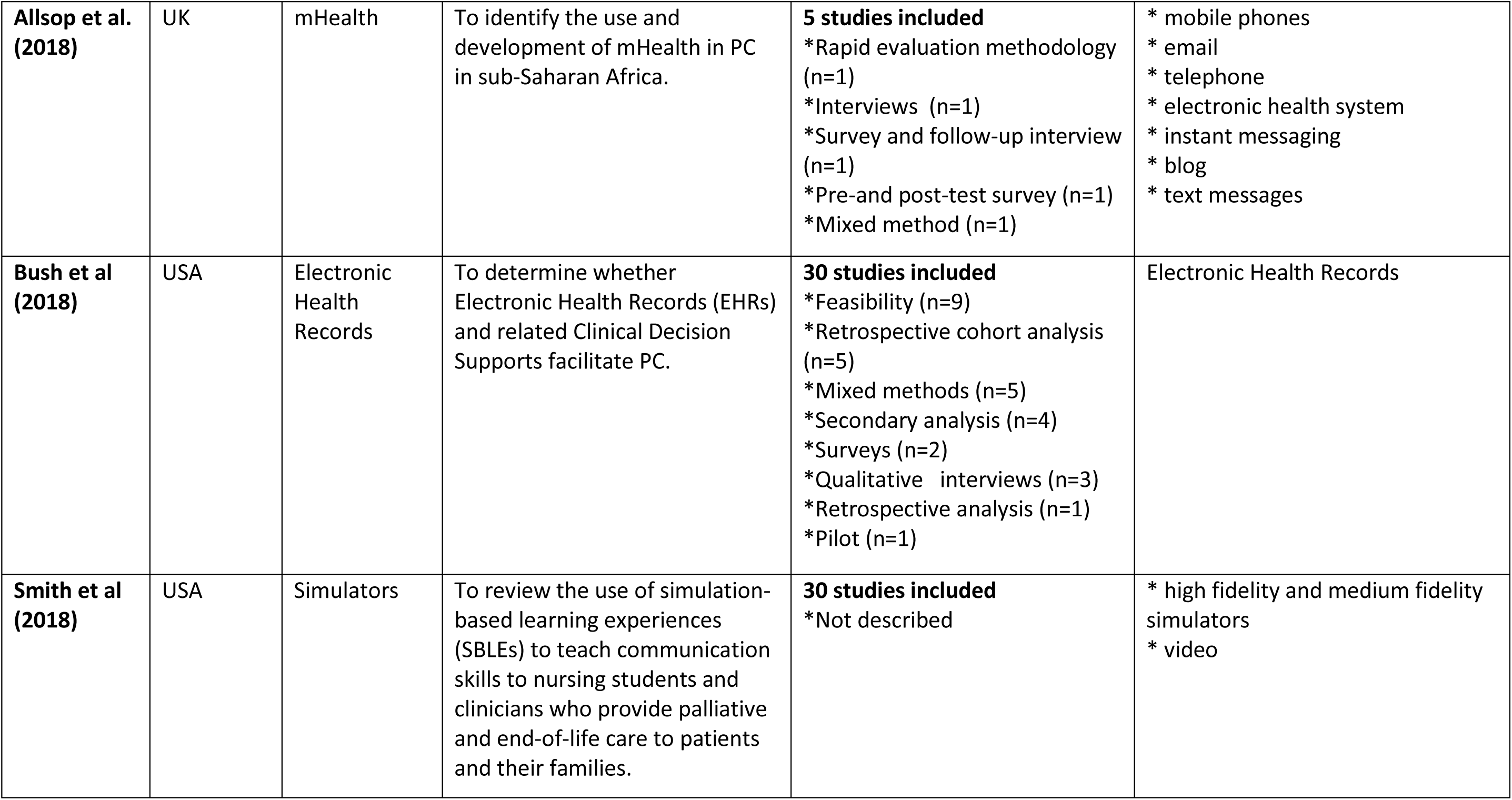
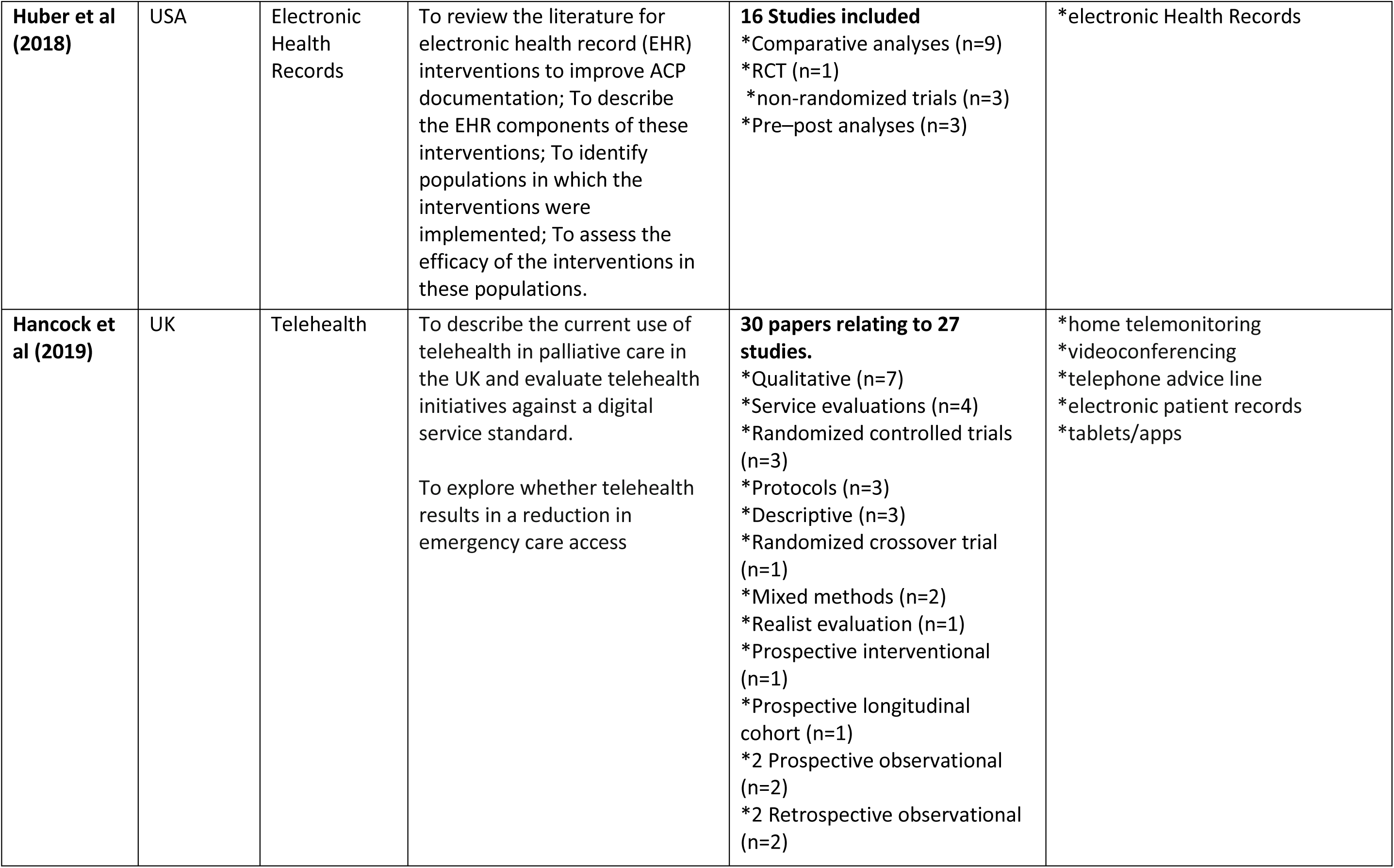
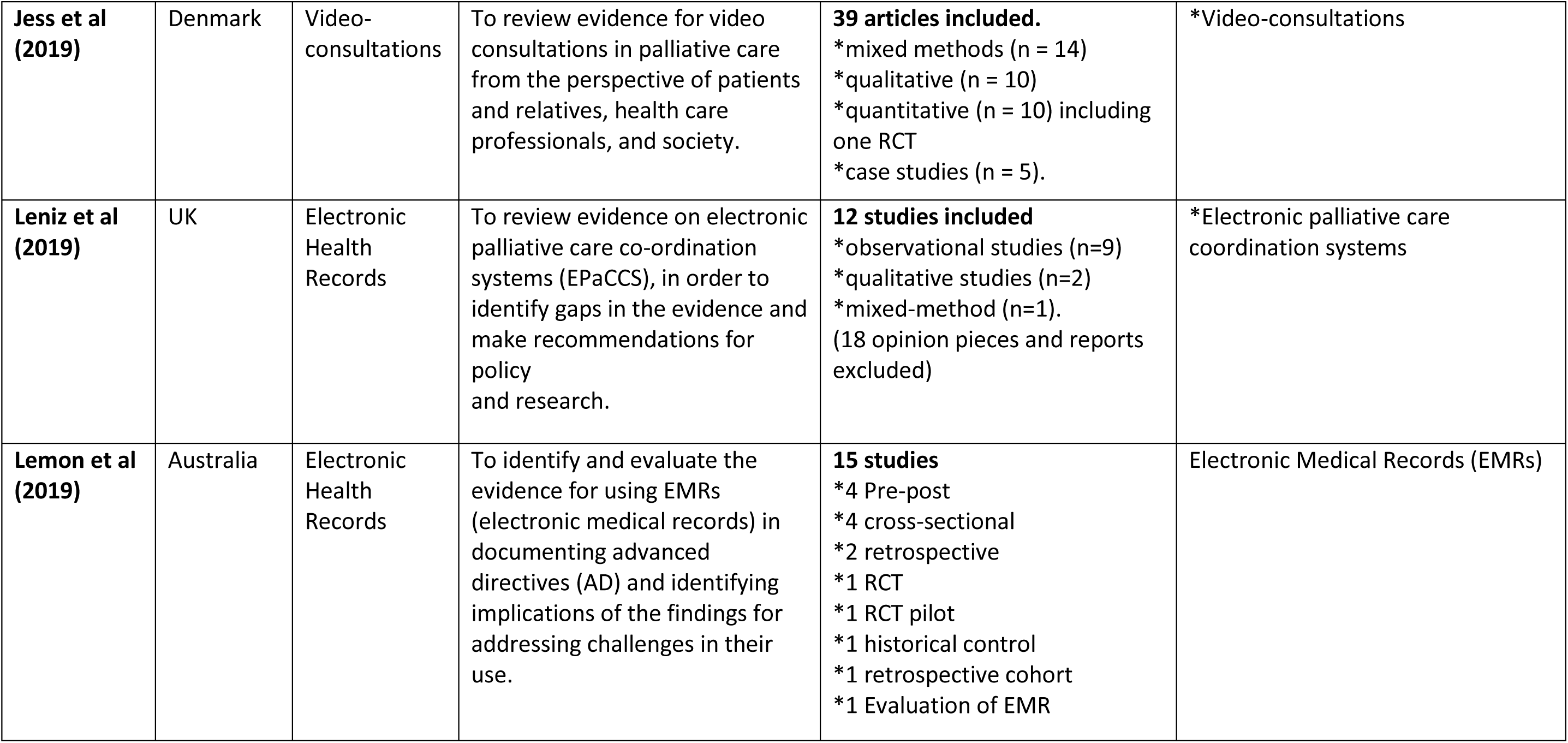
Characteristics of the included systematic reviews (N=21)

The number of studies related to DHIs and palliative care in each review ranged from 5^33^ to 39^37^ Taken together the reviews summarised evidence from 332 unique publications, including four systematic reviews. The 21 reviews were published between 2007 and 2019 and included a total of 41 RCTs. None of the reviews pooled data to perform meta-analyses due to the heterogeneity of included studies.

Ten systematic reviews covered broad areas such as telehealth,^14,16,24,30,31,34^ telehospice,^17^ ehealth^15^ information and communication technologies.^23,25^ Eleven reviews had a more specific focus: EHRs,^27,28,35,36^ internet,^26,38^ weblogs,^39^ mhealth,^33^ telephone,^22^ video conferencing,^37^ and simulators.^29^

#### Types of DHIs described in palliative care reviews

We classified the types of DHIs described in 328 publications included in the 21 reviews (**Table 2**). The most featured DHIs involved videoconferencing or videophone (n = 56, 17%), EHRs (n = 51, 16%) and telephone or mobile phone (n = 41, 13%). Online interventions, including educational websites and online courses, were described in 31 publications (9%). Only six publications were focused on social media (2%), e.g. interactive online blogs. We found a relatively large proportion of studies describing mixed or unspecified DHIs (n = 50, 15%). Mixed DHIs were delivered using a choice of DHIs, or using a DHI with multiple components (e.g. telephone call with follow-up video-consultation). Unspecified studies included general surveys or qualitative studies examining the use of DHIs in general. Studies describing and evaluating EHRs increased over the review period, with more publications on EHRs compared with any other DHI each year since 2016.

**Table 2.** Types of DHIs reported in publications included in 21 reviews (n=328)

| **Type of Digital Health Intervention** | **No. of publications** | **% of publications** |
| --- | --- | --- |
| Videoconferencing/videophone | 56 | 17% |
| Electronic health records | 51 | 16% |
| Phone/mobile phone | 41 | 13% |
| Online | 31 | 9% |
| Video | 27 | 8% |
| Computer | 20 | 6% |
| PDA / tablet /smartphone | 16 | 5% |
| High-fidelity Simulator | 16 | 5% |
| Other (e.g. digital pens) | 7 | 2% |
| Social media | 6 | 2% |
| Telemonitoring | 5 | 2% |
| Text messaging | 2 | 1% |
| Mixed/unspecified | 50 | 15% |
| **TOTAL** | **328** | **100%** |

#### Main purpose of DHIs described in palliative care reviews

DHIs were used for a range of purposes in palliative care **(Table 3)**. A fifth of publications described DHIs for educational purposes (n=64) most frequently involving online learning, simulators and videoconferencing. Symptom management was the main aim of DHIs outlined in 15% of publications, and all types of DHI were used for this purpose. Decision-making support for patients and professionals was the main purpose of DHIs described in 13% of publications - video aids and EHRs were often used for this purpose. Information provision or management, often using EHRs, was the main aim of DHIs in 13% of publications. Communication was the main aim of DHIs in 9% of publications, with videoconferencing most often used. Overall, 15% of publications described DHIs for mixed or unspecified purposes. Mixed purposes could include information support and decision-making; or communication and information-sharing. Unspecified purposes had no specific focus.

**Table 3.**
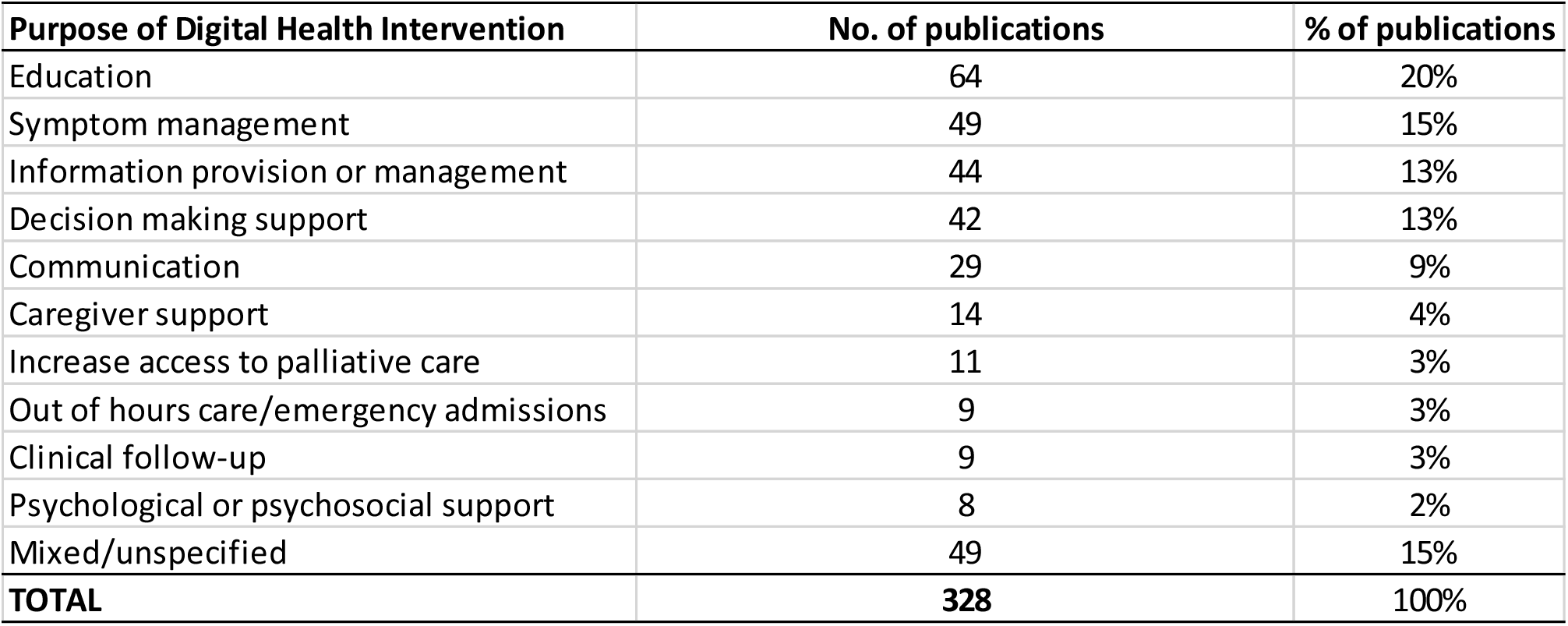
Main purpose of DHIs reported in publications included in 21 reviews (n=328)

| **Purpose of Digital Health Intervention** | **No. of publications** | **% of publications** |
| --- | --- | --- |
| Education | 64 | 20% |
| Symptom management | 49 | 15% |
| Information provision or management | 44 | 13% |
| Decision making support | 42 | 13% |
| Communication | 29 | 9% |
| Caregiver support | 14 | 4% |
| Increase access to palliative care | 11 | 3% |
| Out of hours care/emergency admissions | 9 | 3% |
| Clinical follow-up | 9 | 3% |
| Psychological or psychosocial support | 8 | 2% |
| Mixed/unspecified | 49 | 15% |
| **TOTAL** | **328** | **100%** |

**Table 4.**
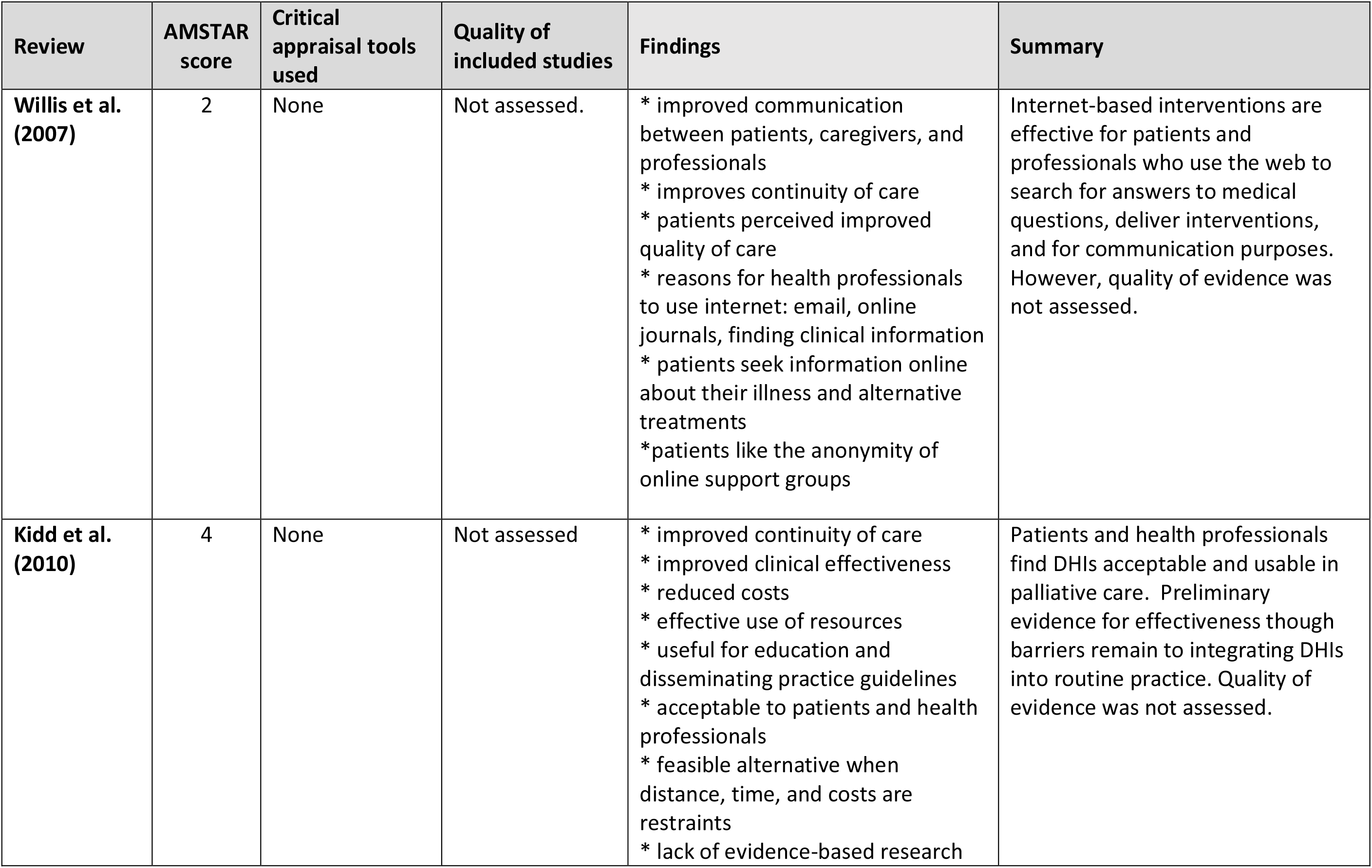
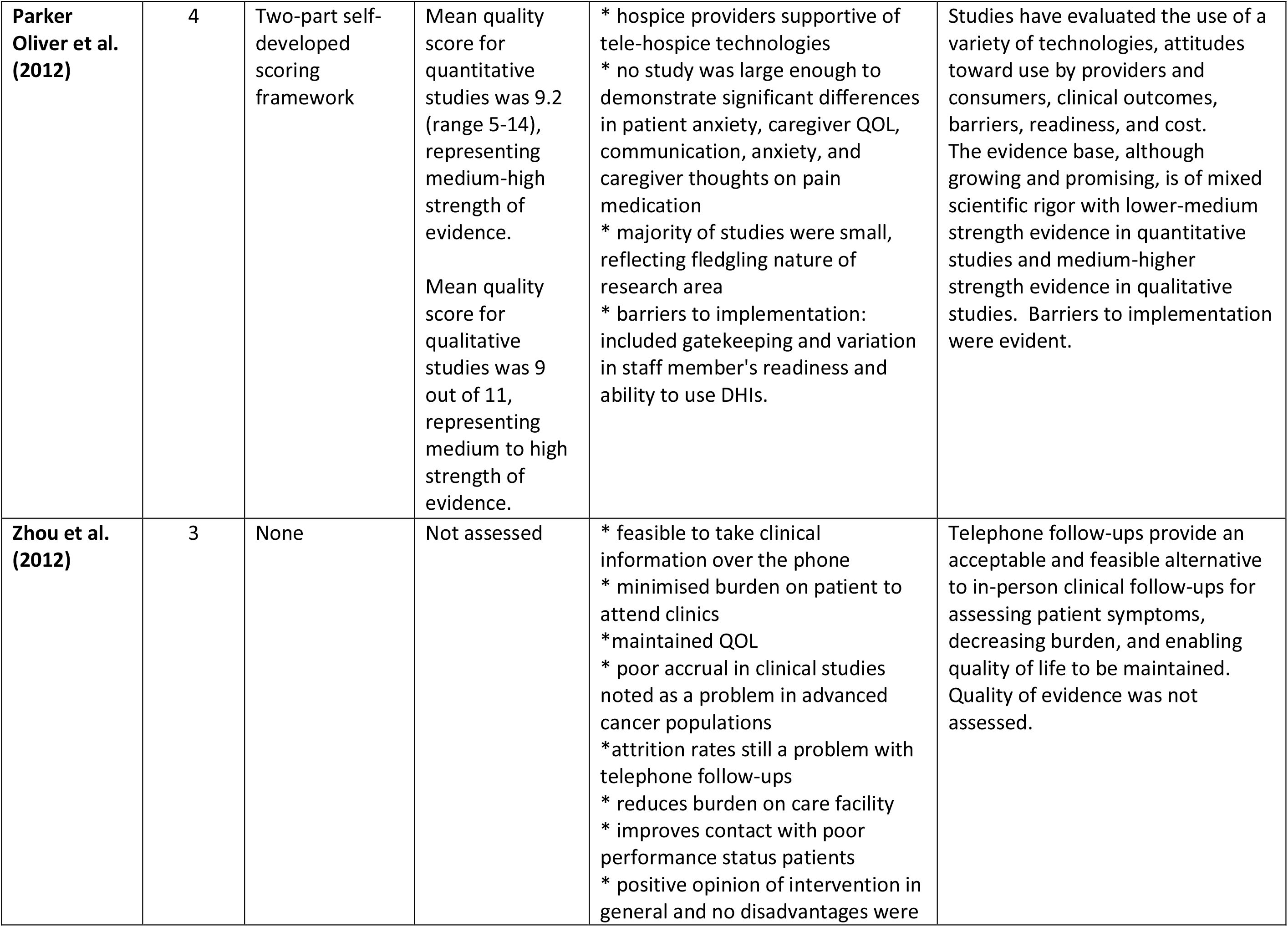
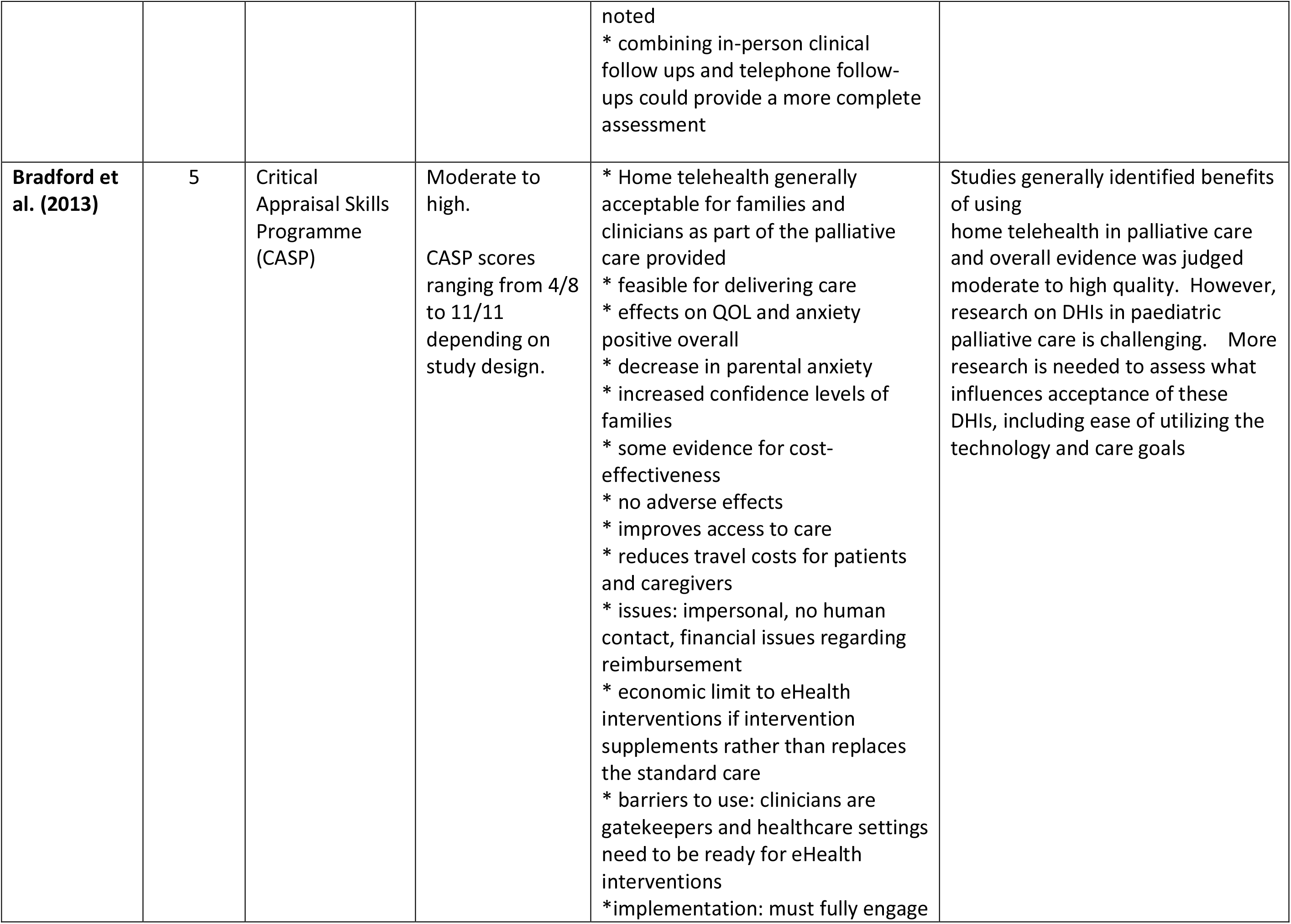
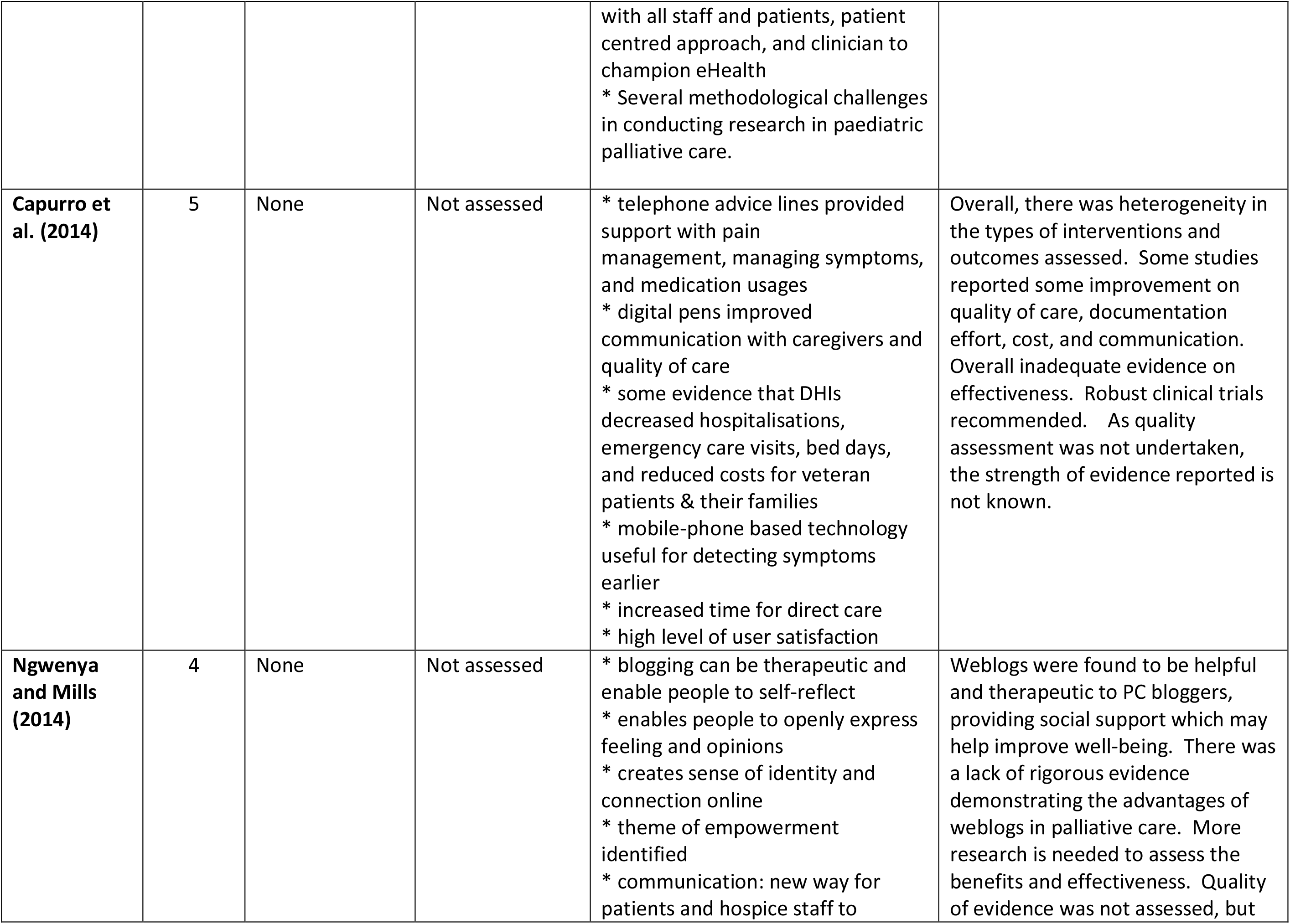
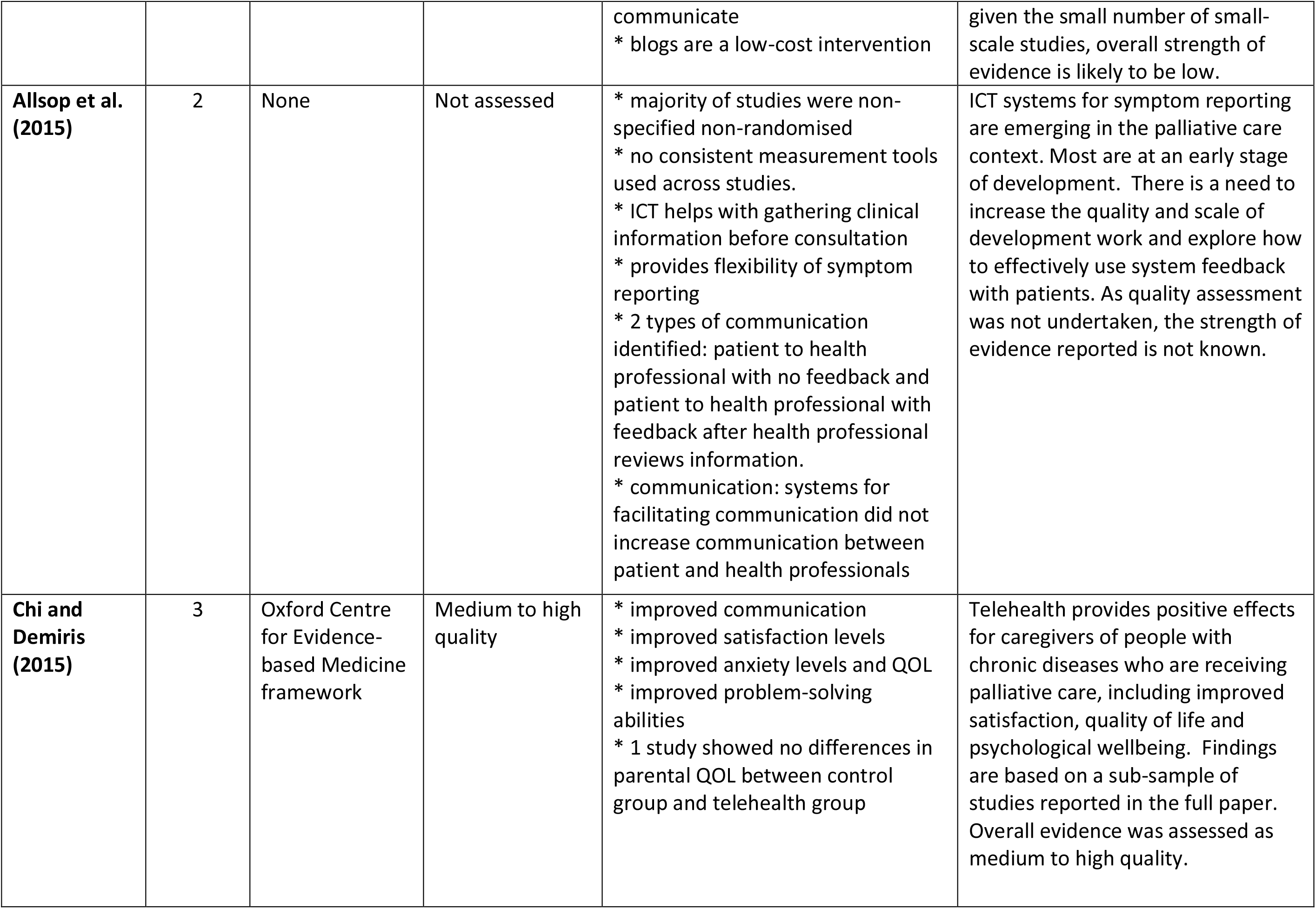
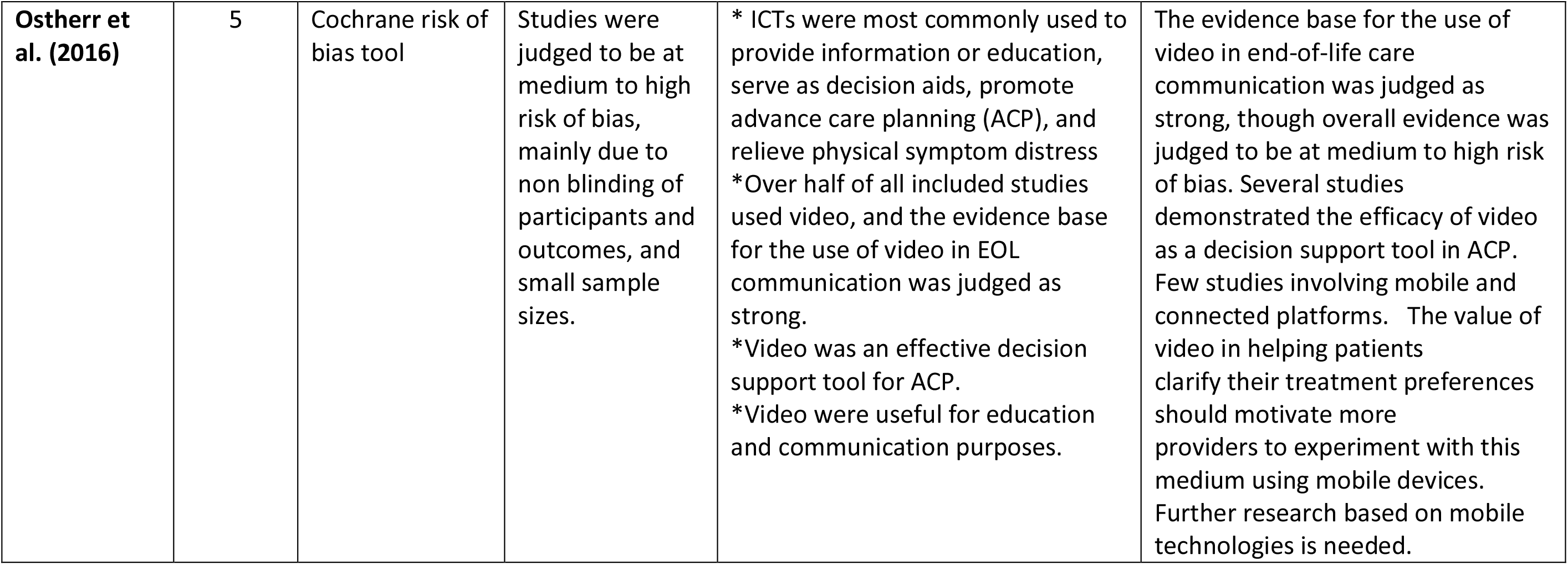
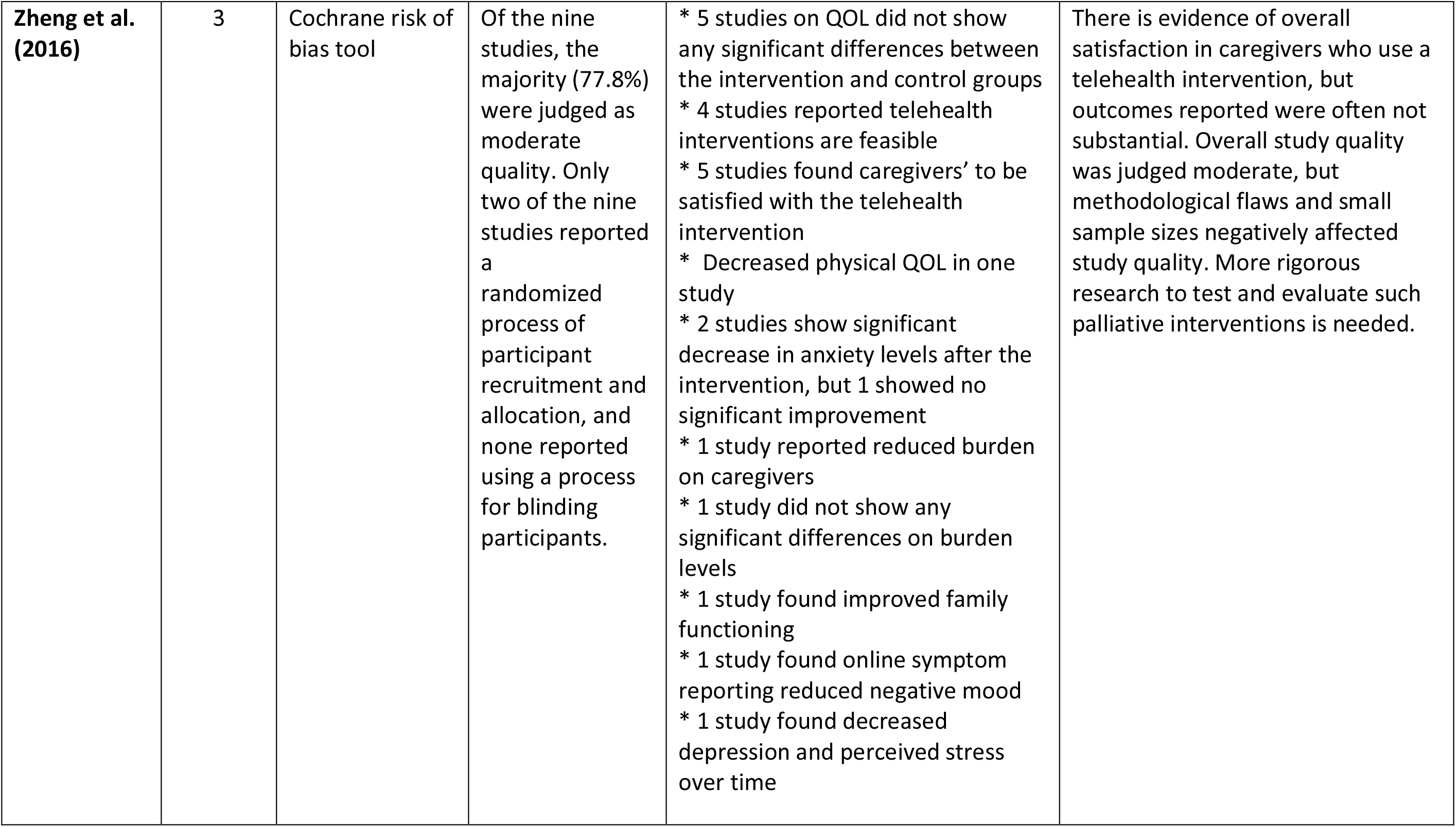
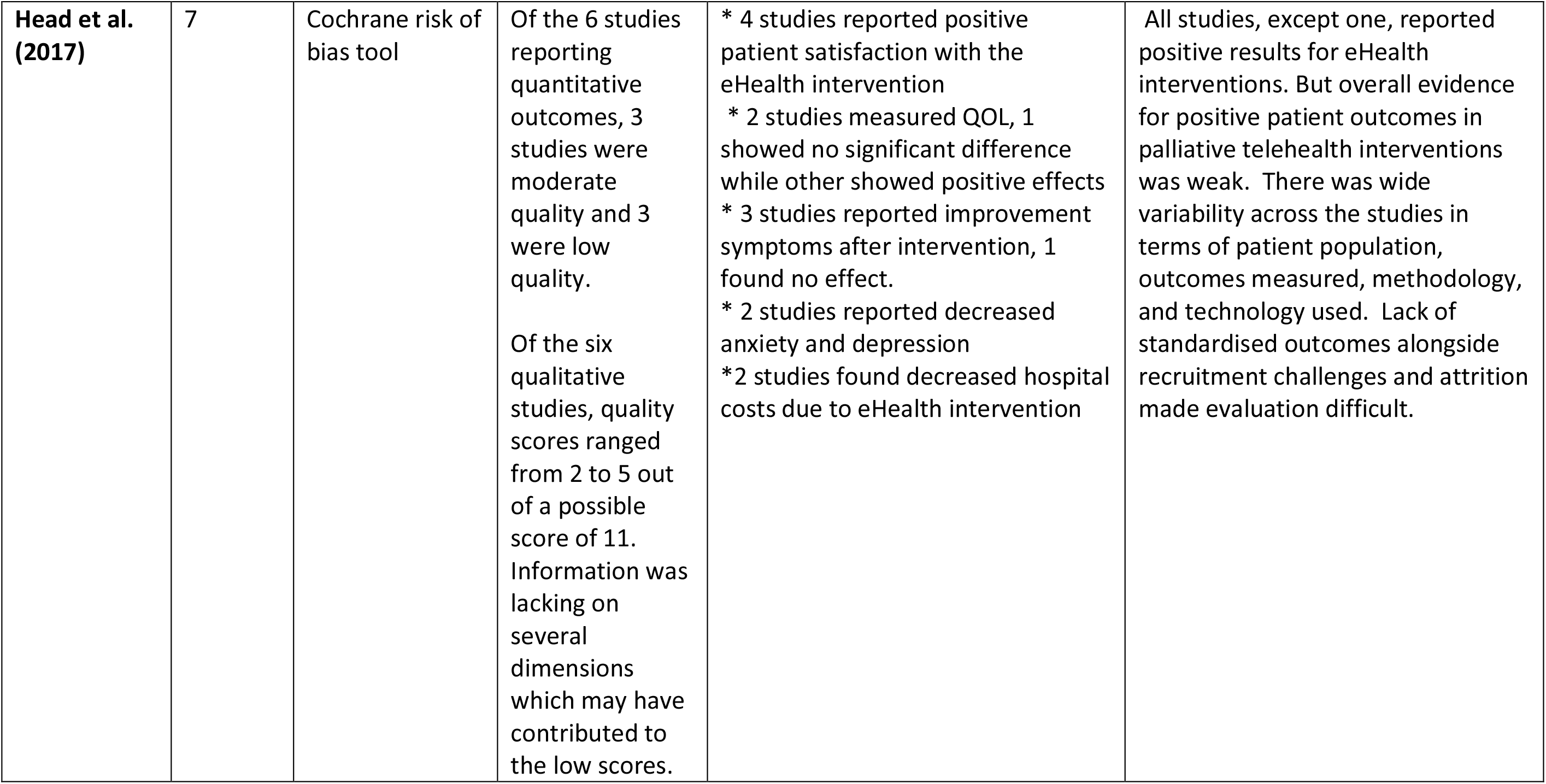
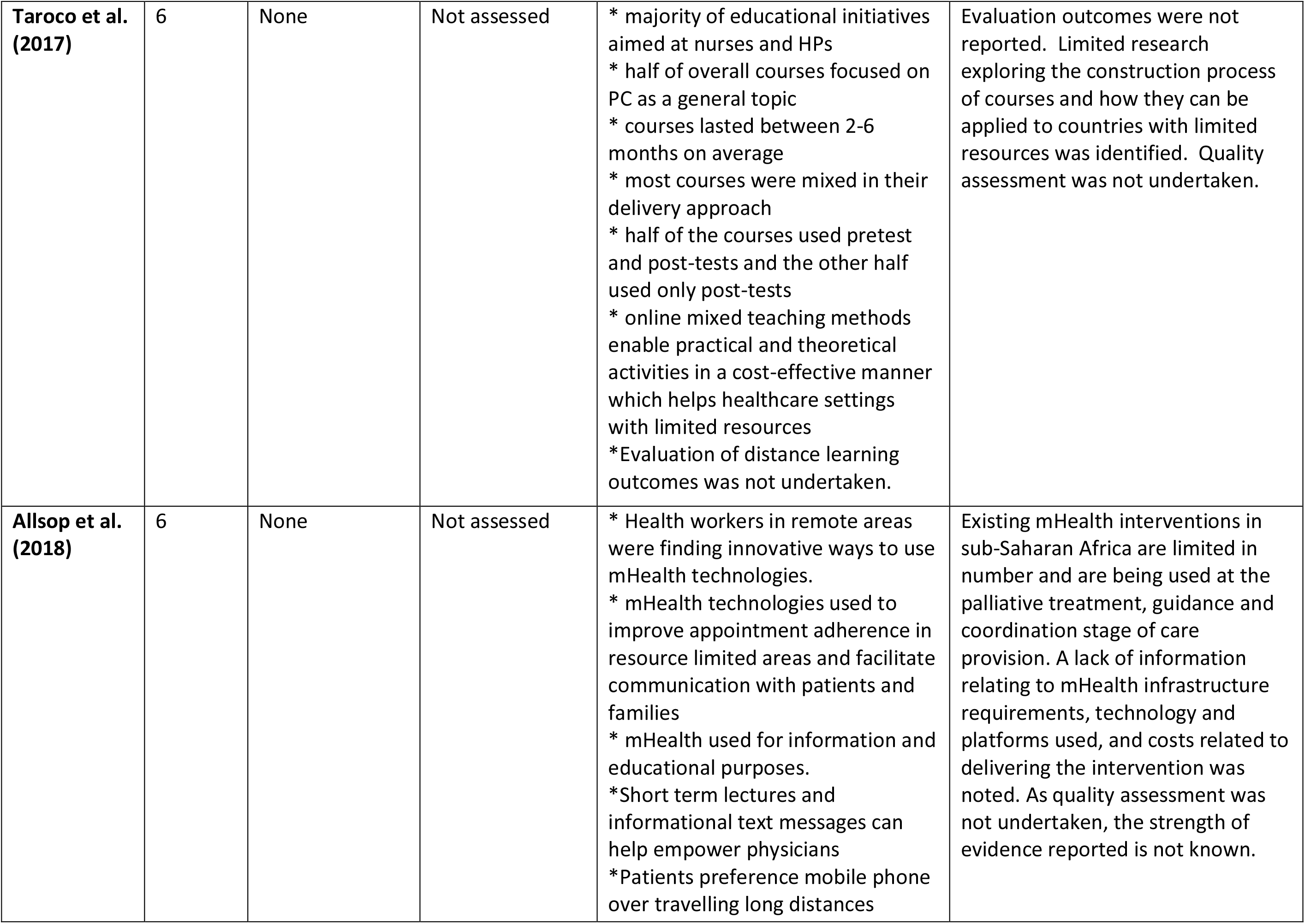
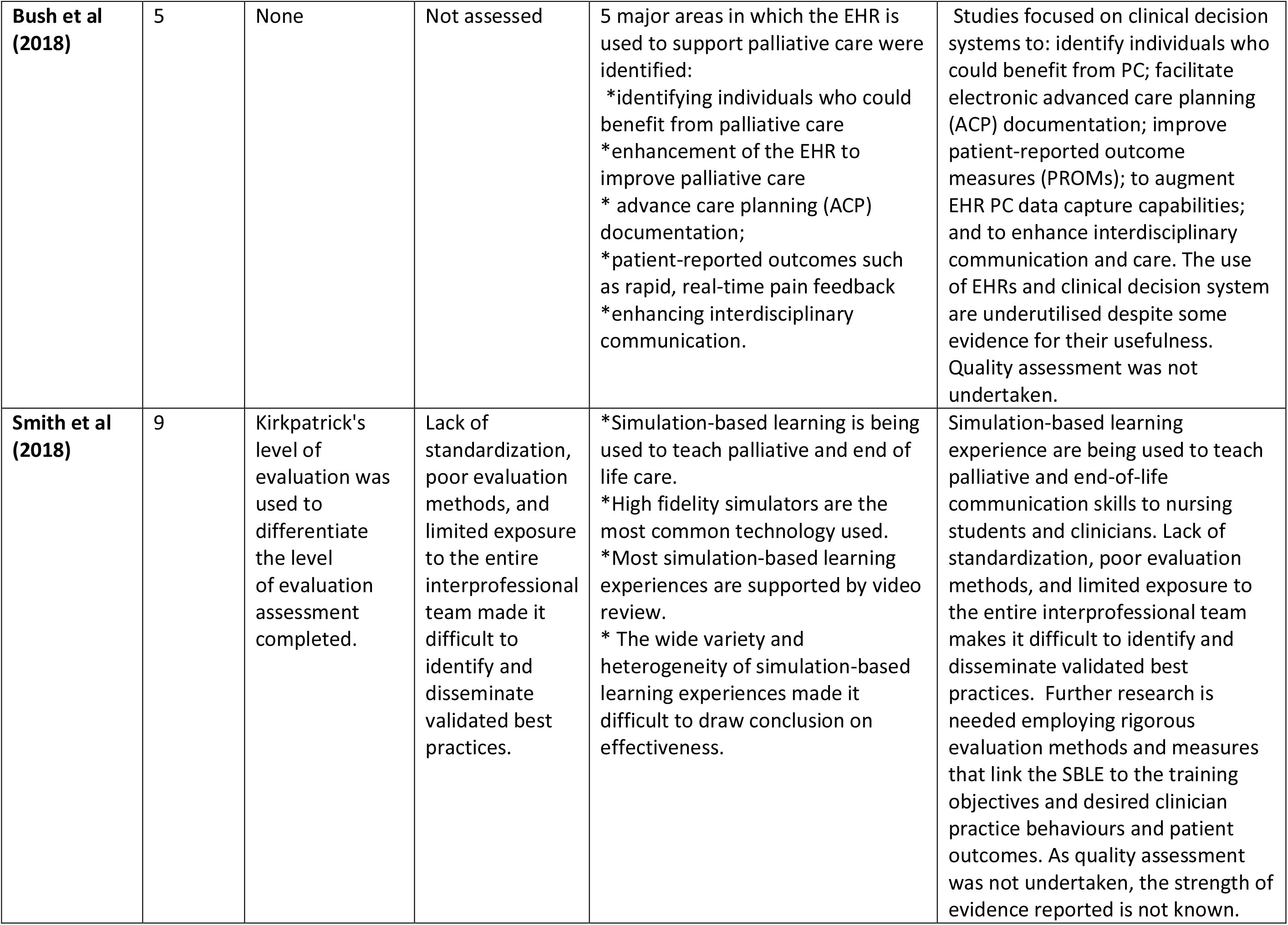
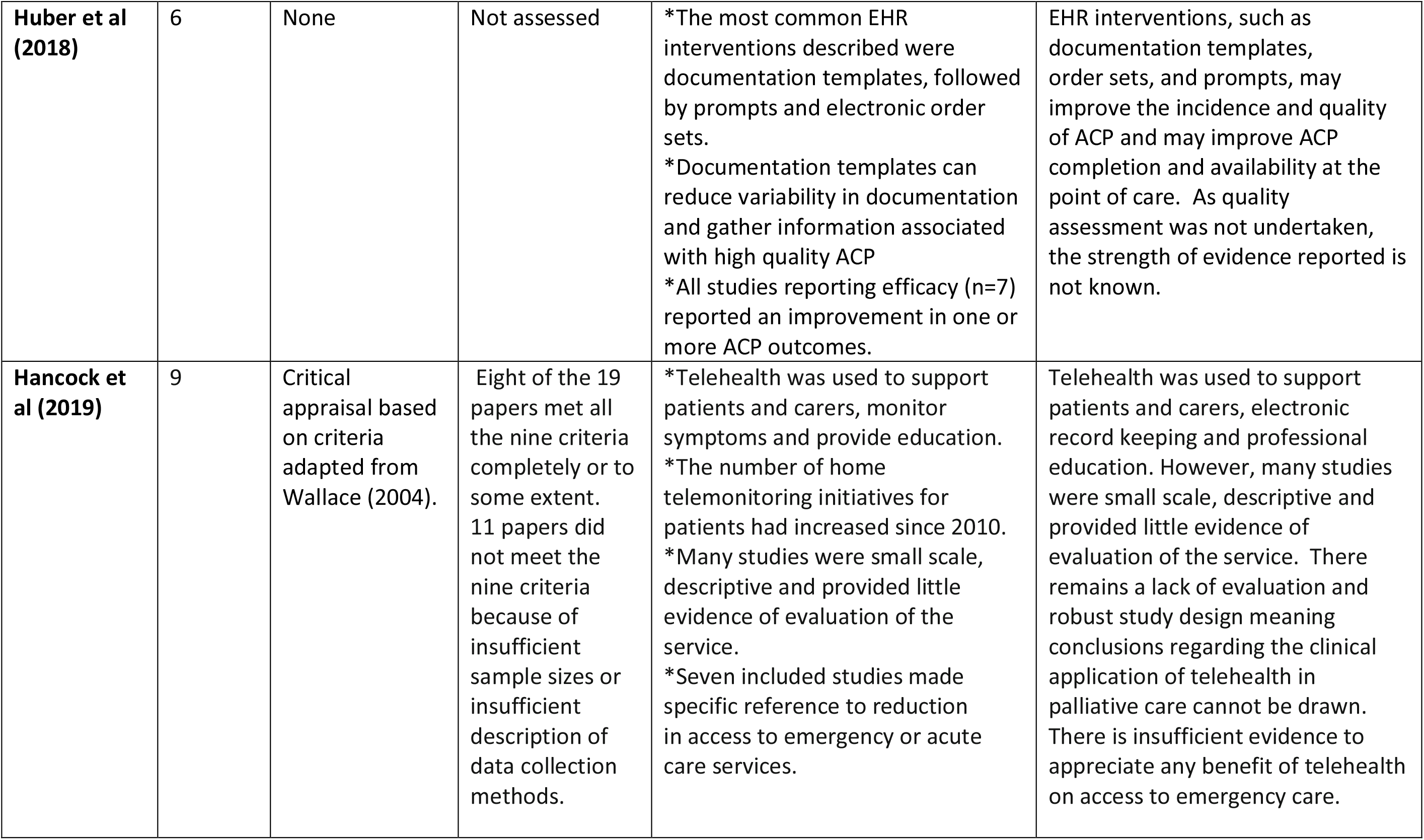
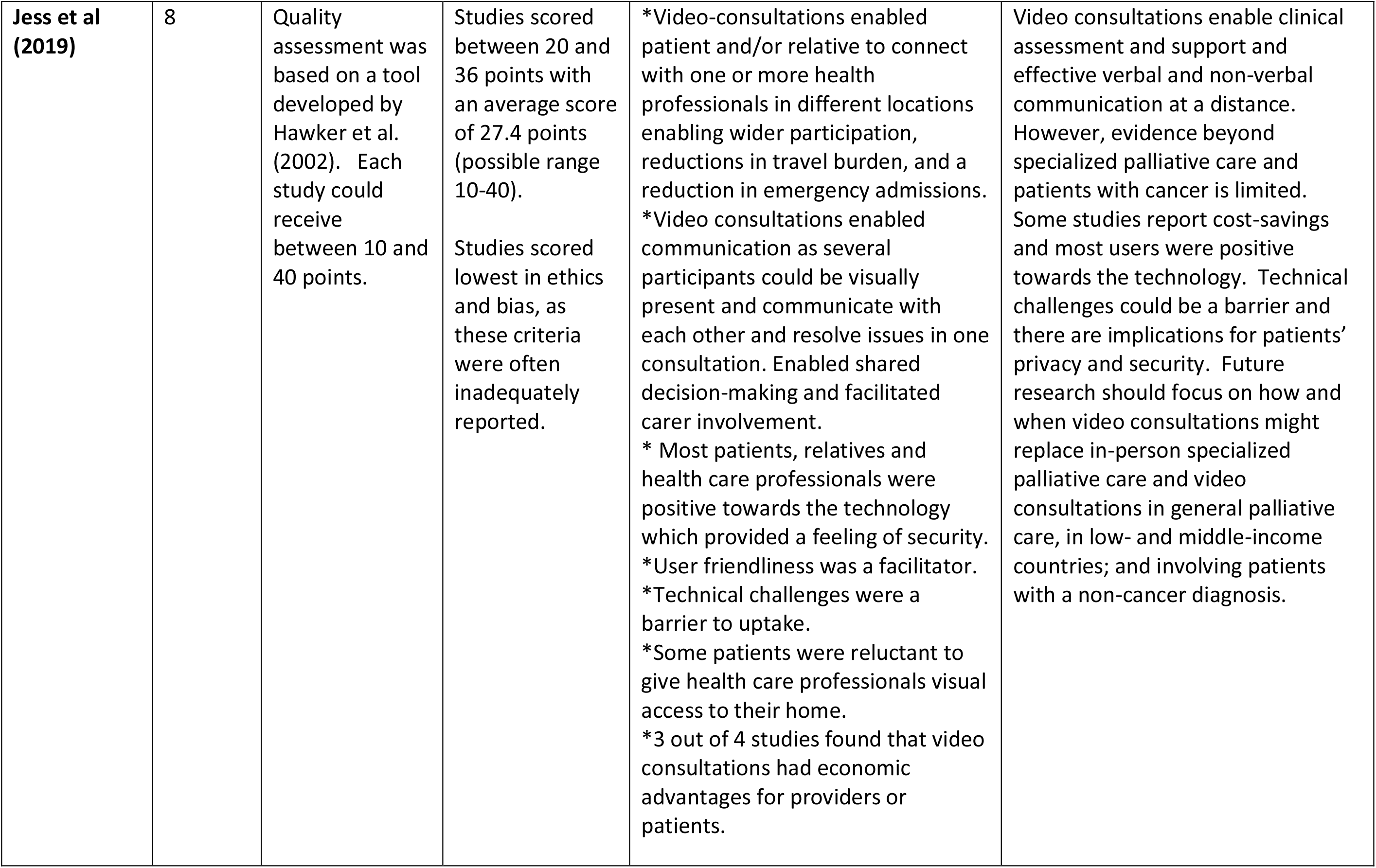
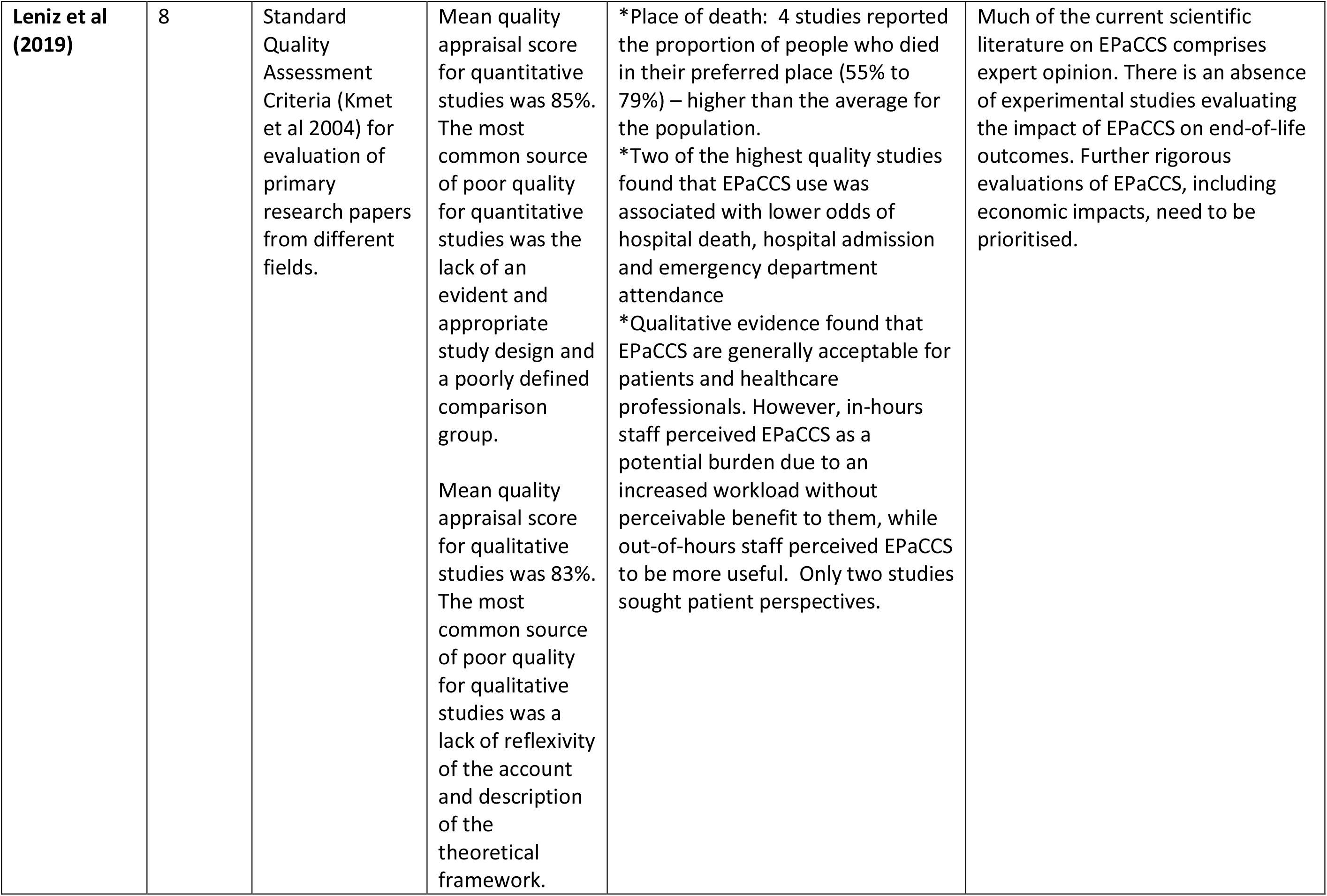
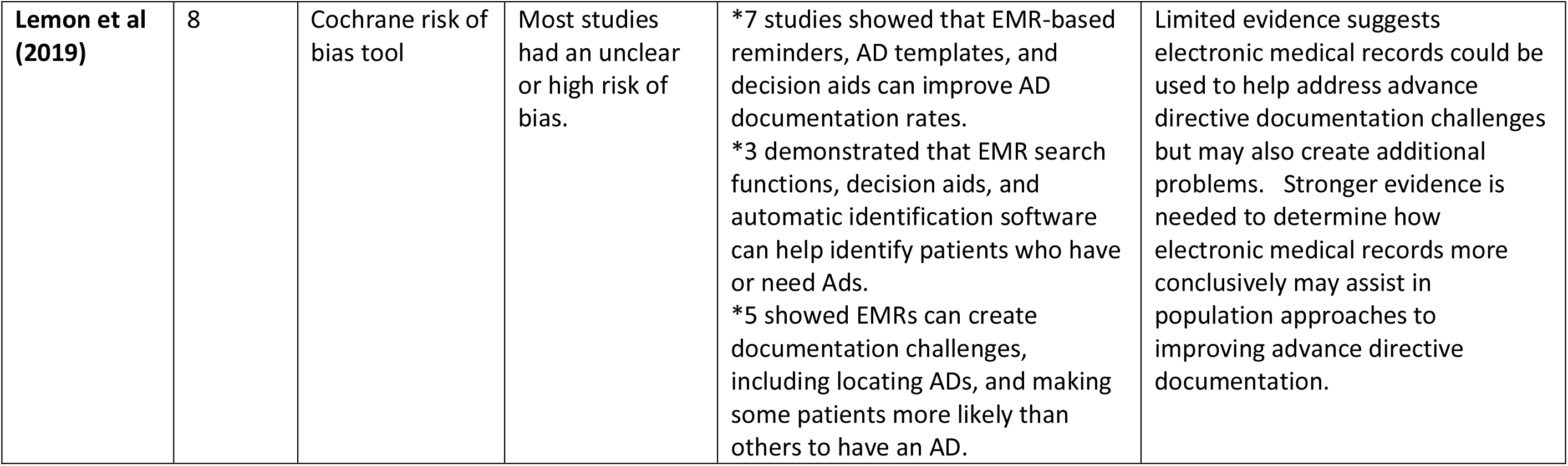
Overall quality and findings from the included systematic reviews (N=21)

### Quality of evidence

#### Quality of the included reviews

The quality of the 21 included reviews varied, with AMSTAR scores ranging from 3 to 10, out of a possible total score of 11 (Supplementary material 1). Most reviews (n=11) were of moderate quality (AMSTAR score between 5 and 8). The median rating of reviews published between 2007 and 2017 was 4, compared to a median rating of 8 for reviews published in 2018 and 2019. All reviews described the characteristics of included studies; most described a comprehensive search strategy (17 of 21) and used independent data extractors (15 of 21). Just over half assessed the quality of evidence and used this information to contextualise the results (11 of 21). Approximately half referred to a protocol or a-priori research objectives (11 of 21). Less than half specified the methods used to combine the results of studies (9 of 21). Only a third of reviews (7 of 21) searched grey or unpublished literature. None of the reviews considered publication bias, and only one included a list of excluded studies.

#### Quality of studies within the reviews

Eleven reviews assessed the quality of evidence.^14,17,24,25,29-31,34-37^ Four used the Cochrane risk of bias tool.^25,30,31,36^ One used the Critical Appraisal Skills (CASP) programme tool.^14^ Five reviews used different tools previously described in the literature^24,29,34,35,37^ while one review developed a quality appraisal framework specifically for their review.^17^ Only three reviews described evidence as moderate to high quality.^14,17,24^ Eight reviews reported evidence of low to moderate quality.^25,29-31,34-37^ This was due to small sample size, insufficient detail on study design, unclear or high risk of bias, non-blinding of participants and outcomes, and poorly defined comparison groups.

## USE AND EFFECTS OF INTERVENTIONS

Findings from each review are described in **Table 4** (See Appendix) in relation to seven thematic areas: education symptom management, information sharing, decision-making, communication, quality of life and cost-effectiveness.

### Education

Eight reviews identified DHIs for education, of which most focused on describing interventions rather than evaluating their outcomes.^15,16,24,25,29,33,34,38^ Educational interventions were delivered via online learning for professionals,^15,25,30,38^ videoconferencing for professionals,^16,34,40^ videos for professionals,^19,25,28^ online symptom reporting for caregivers,^24^ simulation based learning experiences (SBLEs) for professionals;^29^ and mobile phones/text messaging for education and training of providers and patients.^33^ Two reviews reported that online learning was a feasible alternative to in-person training, though quality of evidence was not assessed.^16,38^ In a review of distance learning for healthcare professionals, Taroca et al. suggested that online case consultations involving active participation of students facilitated knowledge retention.^38^ They also noted the prevalence of mixed educational initiatives (i.e. distance learning and classroom based), with 64% of studies involving mixed approaches, suggesting a need for classroom activity to consolidate knowledge acquired at a distance. There was no consensus about the most effective learning methods, and most virtual learning environments used a variety of multimedia to support communication and feedback mechanisms. Kidd et al.^16^ suggested that online learning and remote access to guidelines supports dissemination of good practice but also reported that face-to-face teaching methods are preferred when discussing emotional or psychological issues.^16^ Ostherr et al.^25^ found strong evidence for benefits of video for educating patients about their illness and helping to determine treatment choices, although the evidence was judged to be at high risk of bias. Smith et al. examined evidence on the use of simulation based learning for end-of-life care conversations, though information on outcomes was absent.^29^

### Symptom management

Thirteen reviews referenced the role of DHIs in monitoring, assessing, and managing physical and psychological symptoms.^14-17,22-25,27,30,31,34,37^ EHRs were used to record symptoms^27,34^ while telephone and videoconferencing were frequently used to monitor, assess and treat symptoms.^15,22,23,25,30,37,39^ Some reviews described positive impacts of DHIs on symptom management, while most reviews identified inconsistent evidence or noted that evaluation of impact in many studies was lacking. In their review of telephone follow-up for patients with advanced cancer, Zhou et al.^22^ concluded that telephone follow-up is a feasible alternative to hospital follow-up for symptom palliation, and reduces travel burden. Head et al.^31^ reported positive or no impacts of DHIs on patient symptoms (e.g. physical and social functioning), noting that overall evidence was weak. Jess et al.^37^ described positive impacts of videoconferencing on symptom burden, especially in remote settings, though also noted negative impacts in a some studies specifically due to technical challenges which frustrated communication.^37^ Hancock et al.^34^ described home telemonitoring initiatives for patients (e.g. use of the telephone or computer software to record clinical symptoms at home); however most interventions had not been evaluated. Similarly Kidd et al.^16^ described uses of telephone hotlines and electronic questionnaires to inform symptom management, though there was little evaluation. The heterogeneity of outcomes used to assess particular symptoms such as pain was highlighted by Allsop et al.^23^ Bush et al.^27^ described evidence linking the documentation of clinical symptoms on an EHR to reduced time in hospital in the last 6 month of life; though this finding was based on just one of the studies included in their review, as it was not directly evaluated in others.

Seven reviews reported effects of DHIs on psychological symptoms - anxiety, depression and distress.^14,17,24,30,31,37,39^ Bradford et al.^14^ described a number of small studies examining videoconferencing interventions for paediatric palliative care, noting reductions in anxiety. Chi et al.^24^ found enhanced psychological health in caregivers (less anxiety, depression, stress, burden, irritation and isolation) associated with DHIs. Similarly Head et al.^31^ identified positive effects of DHIs (telemonitoring and videoconferencing) on patient anxiety, depression and distress. Zheng et al.^30^ reported significant improvements in caregiver anxiety associated with access to videophones. Oliver et al.^17^ identified studies examining the effect of DHIs on anxiety, though studies were not large enough to detect significant differences in outcomes. Jess et al.^37^ found mostly positive impacts of video-conferencing on patient and caregiver anxiety, with the exception of one RCT which found negative impacts.^41^ This RCT compared weekly video-consultations by a palliative care specialist with treatment as usual in home-dwelling patients with advanced cancer. The authors concluded that higher distress in the video-consultation arm may have been due to excess focus on symptoms and suffering, and the provision of pre-scheduled support over 3 months as opposed to when it was actually needed.^41^ Ngwenya et al.^39^ focusing on online blogging, reported that patients experienced a sense of emotional support, social connections and empowerment through writing online blogs.

### Information sharing

Nine reviews considered the information-sharing value of DHIs, with most describing the value of the information rather than evaluating specific outcomes.^15,16,23,26-28,35,36^ In an early low quality review of internet use, Willis et al. described the positive impacts of the internet as an additional source of information for patients, families and clinicians.^26^ They found that patients and carers used online support groups and chatrooms to exchange information and support about an illness and alternative treatments. Patients and carers developed a connection with others online and appreciated the anonymity associated with online support. Capurro et al.^15^ reported that DHIs were used by clinicians, patients and carers to meet informational needs regarding pain and symptom management and medication use; although the quality of evidence was not assessed. Kidd et al. highlighted the importance of telephone helplines for general practitioners (GPs), nurses, and caregivers for gathering information about managing symptoms and medical equipment.^16^ These telehealth interventions improved the reliability and accuracy of information exchanged.^16^ Allsop et al.^23^ noted that many systems designed to capture information from a patient for use by a healthcare professional, involved relaying symptoms without engaging in active forms of communication.

Four reviews highlighted the information-sharing function of EHRs in palliative care.^27,28,35,36^ All were moderate quality. These reviews concluded that EHRs available across settings and platforms allow patient preferences regarding advance care planning (ACP) to be shared; improving continuity of care and ensuring that patients are treated in line with their wishes. Bush et al.^27^ reported that in low resource settings, the implementation of a standalone EHR system capturing patient demographics and palliative care treatment information was found to significantly improve clinical workflow. Leniz et al.^35^ found that those with an EHR shared across settings were more likely to die in their preferred place compared with those who did not have an EHR. However, EHRs were limited in their capacity to capture important qualitative information such as information on anxiety or family distress.^27^ Furthermore, finding the location of relevant ACP information within the EHR was often challenging;^36^ though could be improved by ensuring all ACP information is documented in a specific area.^27^ Documentation templates, order sets and prompts, may also improve the quality and incidence of ACP within EHRs.^28^ Having an EHR improves documentation of advance care plans and communication of care planning information;^27,28,36^ but this can come at the cost of increased workload,^35^ challenges identifying which patients should have a shared EHR,^36^ and concerns regarding data-sharing, security and consent.^35^ Huber et al.^28^ suggest that further research focused on developing a consensus definition for ACP documentation and related quality elements in EHRs is needed.

### Decision-making

Four reviews considered the role of DHIs in decision-making by patients^25^ and professionals.^27,36,38^ Ostherr et al. identified 20 studies where video, computer-based multimedia and online materials were used as patient decision-aids to support.^25^ There was strong evidence for the efficacy of video in facilitating ACP decisions, resulting in improvements in completion of advance directives, discussion of end-of-life preferences, and improved patient knowledge and satisfaction. Taroca et al. identified two studies on distance learning courses for decision-making in palliative care, but did not describe the outcomes.^38^ Two reviews considered the role of clinical decision support systems (CDS), including EHRs in facilitating decision-making.^27,36^ Bush et al.^27^ described evidence on the use of such systems to identify patients for a palliative care approach, and to capture ACP directives and patient reported outcomes to inform clinical decision-making. Due to heterogeneity of studies, evidence could not be synthesized, but Bush et al.^27^ described positive impacts including a reduced likelihood of ICU admissions and hospital death for those with patient reported outcomes shared via EHR, compared to those without; and earlier identification of patients for ACP discussion. Lemon et al.^36^ found that EHRs can improve documentation of advance directives. Electronic reminders, electronic templates, decision aids and standard locations of advance directives can increase documentation of advance directives. Electronic search systems and identification algorithms located within the EHR can assist with identification of patients who could potentially benefit from a palliative care approach, by flagging those who may have palliative care needs for review by the clinician. Overall, evidence described by Lemon et al. was weak, but points towards promising effects of EHRs for ACP.

### Communication

Ten reviews described the role of DHIs to facilitate communication between patients, professionals and carers using phones, internet, and computer systems.^15,23-26,29,31,33,37,39^ Positive effects included enhanced communication between patients, healthcare professionals and caregivers;^15,24,26,37^ more opportunities to express feelings;^39^ increased connectednesss;^15^ caregiver support^17^ and improved ACP.^25^ Jess et al.^37^ identified 16 studies relating to the impact of videoconferencing on communication in palliative care. Positive impacts included greater efficiency and access, whereby several participants could be visually present and participate at once; shared decision-making involving the multidisciplinary team, patient and family; and enhanced communication through access to non-verbal as well as verbal responses. Negative impacts could occur where the family felt overwhelmed by the involvement of too many participants. Smith et al.^29^ found that simulation based-based learning was frequently used to teach nursing students communication skills in palliative care settings, but due to the lack of standardization and poor evaluation, it was difficult to identify best practices.

### Quality of Life

Seven reviews considered the effects of DHIs on quality of life (QoL).^14,17,22,24,30,31,37^ Most reviews described improvements that were not statistically significant or positive impacts; negative impacts were rarely observed. Zheng et al. found no significant difference in QoL outcomes after telehealth interventions for caregivers.^30^ Head et al. identified one study reporting a positive impact of telephone monitoring on QoL whereas another involving videophones showed no difference.^31^ Similarly, in their review of telehealth for paediatric palliative care, Bradford et al^14^ found either positive effects on QoL or no significant differences. Zhou et al. reported that telephone follow-ups with patients with advanced cancer reduced the patient burden by eliminating the need to come into hospital, facilitating a better QoL, though quality of evidence was not assessed and insufficient data on included studies was provided.^22^ In a review of telehealth and hospice care, Oliver et al.^17^ reported that studies examining QoL were too small to identify clinically significant differences.

In their review of videoconferencing, Jess et al. identified several studies incorporating a QoL measure in their design, but QoL outcomes were not described in their key findings.^37^ In a review of weblogs in palliative care, Ngwenya and Mills^39^ concluded that weblogs improve patient and QoL by empowering patients and giving them a sense of active participation in their treatment, but this was a small scale study with no quality assessment of included studies. In reviews of EHRs, outcomes relating to QoL were rarely assessed.^34,35^

### Cost effectiveness

Five reviews considered the financial implications of DHIs, with most reporting positive impacts of DHIs on costs.^14-17,37^ Jess et al. described cost savings associated with video-consultation in palliative care for clinicians, service providers, patients, and caregivers.^37^ In two studies included in their review, video consultations between healthcare professionals and patients resulted in cost-savings for the hospital, compared to in-person consultations, and in clinician travel expenses for home visits. Travel cost savings were also noted for patients and carers in rural settings.^37^ In a review of DHIs in hospices, Oliver et al.^17^ identified one telehospice cost analysis study; this study reported reduced costs for telehospice visits versus traditional hospice homecare. Bradford et al.^14^ described cost efficiencies when video-visits were used in place of home visits; and when videoconferencing was used to educate patients about self-care; but cautioned that the cost-effectiveness will depend on whether DHIs are used in parallel with, or as a replacement for, traditional approaches. Kidd et al. described DHIs as an efficient alternative for patients and clinicians when time and distance is limiting.^16^ Capurro et al. described cost efficiencies related to reduced hospital visits, but this was based on only one study in their review.^15^ Overall, evidence on cost effectiveness was positive, though interventions and outcomes assessed were heterogeneous, findings were based on a small number of studies, evidence quality was not always assessed and robust economic evaluation not undertaken.

## DISCUSSION

### Main findings

The evidence captured in this meta-review indicates that DHIs in palliative care are being used for education, symptom management, information-sharing, decision-making and communication, with the aim of improving patients’ quality of life and the reach and efficiency of services. The methodological quality of the included systematic reviews was mostly moderate, although those published since 2018 tended to be stronger. Primary studies appraised in the reviews were typically of low to moderate quality. Positive impacts of DHIs were reported on education, information-sharing, decision-making and communication in palliative care contexts.

Mostly positive effects, or no negative effects, were noted for psychological symptoms and quality of life. For physical symptom management, evidence was inconsistent or absent. No evidence of risks to patient safety was reported. Systematic review authors conclude that DHIs can play a positive, enabling role in palliative care but call for more rigorous evaluation, implementation, and cost-effectiveness studies, with a greater focus on patient perspectives.

### Advantages of this study

To date this is the most comprehensive meta-review focused on DHIs in palliative care. Compared to a previous meta-review which encompassed six reviews,^20^ it examined a wider range of databases and identified 21 systematic reviews for critical appraisal and synthesis. This meta-review shows that DHIs are more prevalent in palliative care than previously described; are used for a broader range of purposes, that impacts are generally positive, and overall quality of research evidence is improving.

### Limitations of this study

The heterogeneity of reviews aims, methods, and presentation of results created challenges for evidence synthesis. In many reviews DHIs were described but outcomes were not described or evaluated in any detail. Although the searches were completed in January 2020, the dates of the primary studies ranged from 1997 to 2018, reflecting the time lag in academic publishing. None of the eligible systematic reviews focused on smartphone applications for palliative care, despite their growing use in this context.^42,43^ Two reviews emerged after our searches had been completed, including a rapid review on video-consultations in palliative care in context of COVID-19,^44^ and a scoping review of patient experiences of telehealth for palliative care at home.^45^ Neither would have been eligible, as they were not systematic reviews, however we suggest that future meta-reviews include all review types.

### Methodological gaps

The meta-review findings echo the wider literature on digital health^46^ and palliative care,^47^ which point to the need for more rigorous evaluations, cost-effectiveness analyses, implementation studies and patient centred research. The lack of rigorous cost-effectiveness studies seen in the literature on DHI in palliative care, reflects findings from previous metareviews^48,49^ and systematic reviews^50-52^ in digital health. There is a need for greater clarity on what is being compared in cost-effectiveness studies, and whether the DHI is offered in addition to, or as a replacement to the standard approach.^14,25,37^ Undertaking large, well-powered RCTs on DHIs is challenging, partly because technological developments may outpace the timescale for conventional clinical trials;^53^ and also because, in practice, DHIs are implemented in complex systems as opposed to controlled settings.^54^ More rapid research paradigms,^53^ using responsive pragmatic designs that take account of the context and setting in which the DHI is being evaluated and pay greater attention to the factors that facilitate or hinder adoption, may be more realistic and fruitful in future evaluations of DHIs for palliative care. Interdisciplinary evaluation, combining economic, social and clinical research, is needed to better understand the role of different settings, healthcare needs and patient preferences for ensuring the appropriate, safe, acceptable and sustainable use of DHIs in palliative care. Early user involvement (patients, caregivers and staff) will also be key in the design, evaluation and implementation of DHIs in this setting.^55^

### Technology evidence gaps

Personal health monitoring devices, such as wrist-worn activity trackers and smartwatches are now widely used and have been evaluated in other digital health contexts.^56^ The absence of evidence about the use of these may reflect the fact that most studies of trackers are taking place in the context of chronic disease management. Nevertheless, it suggests a need for further research in palliative care, particularly for patients managing at home, for whom wearables and ambient computing (e.g. smart homes) are likely to be increasingly useful. The included systematic reviews did not include studies on the use of smartphone apps. Descriptive reviews on the potential that such apps may have in palliative care are emerging and further research is warranted.^42,43^ Studies using machine learning and artificial intelligence for risk detection and prediction, or for delivering personalised support based on data from individual patients, were also not represented amongst the included reviews, despite progress in AI-enabled healthcare delivery.^57^ Research exploring the use of machine learning using EHRs to predict mortality, and identify patients who would benefit from palliative care shows promise; future reviews need to consider this emerging evidence.^58^ Studies involving robots or chatbots were not identified despite their potential application in palliative care.^59^ Evidence on these types of DHIs in palliative care is needed, to understand their benefits and risks.

### Stakeholder evidence gaps

The WHO has developed a classification framework for DHIs which provides a shared vocabulary for all stakeholders, including researchers, when evaluating effectiveness and identifying gaps in the implementation of DHIs across healthcare settings.^60^ The WHO organizes DHIs into overarching categories by user group: clients (e.g. patients or carers), healthcare providers, health system or resource managers and data services. Most of the research evidence on DHIs in palliative care identified in this metareview was focused on DHIs for healthcare providers (e.g. healthcare provider decision support, remote consultations; healthcare provider communication and training) and to a lesser extent for clients/ patients (e.g. client-to-client communication via online peer group support). No research on interventions for health system managers or administrators in palliative care was found. Using the WHO framework to situate research on DHIs in palliative care and identify gaps facilitates engagement with the wider health and social care sector and highlights the type of DHIs that may need to be prioritised for development and evaluation.

### Telemedicine and related evidence gaps

Most of the evidence identified in this meta-review focused on telemedicine, specifically remote consultations via phone and video. This evidence is timely as the Covid-19 pandemic has pivoted attention towards these approaches.^61^ This meta-review found that remote consultations are feasible in palliative care and generally acceptable to patients^14,16,22,37^ and caregivers.^30,37^ Remote consultations are perceived as particularly helpful when increasing access to care for families who are otherwise isolated by geography or housebound,^14^ reflecting the context for many patients and families due to social distancing requirements during the COVID pandemic. This should help reassure healthcare professionals that patients and caregivers often welcome these approaches, especially when face-to-face options are limited.

While guidance regarding undertaking a remote consultation in palliative care is emerging,^62^evidence gaps remain. There is a need for research to determine when a face-to-face consultation is essential for terminally ill patients and when remote consultation is sufficient or preferred. Research is needed to understand contextual factors influencing the acceptability or effectiveness of remote consultations in palliative care^49^ and to shed light on inconsistent findings around symptom management.^63^ Critically, research on equitable access to palliative care delivered using DHIs is urgently needed to ensure that all those who need palliative care can benefit from it.

### Palliative care research participation

Research involving people who are terminally ill is difficult due to the perceived vulnerability of the population and professional caution.^40^ Professional gatekeeping is a challenge,^64^ and biased samples consisting of patients who are mostly well or particularly motivated is often problematic. However there is ample evidence that many terminally ill patients are interested in taking part in research and may benefit from doing so.^65,66^ As patients and caregivers grow accustomed to receiving care remotely, there will be more opportunities to engage patients and their families in research remotely, reducing burden and travel costs.

Providing a variety of ways in which patient and caregiver data can be collected, including online interviews and focus groups, maximises research participation, and is recommended.

## Conclusions

DHIs are increasingly being implemented in the context of palliative care and the Covid-19 crisis has given this further impetus, particularly for clinical and supportive interventions at a distance. This meta-review has synthesised the corpus of research evidence represented by existing systematic reviews in this area. Overall, this indicates that DHIs are can be useful, safe, and acceptable to many terminally ill patients, their caregivers and staff involved in their care. DHIs are frequently used for education, symptom management, information sharing, decision-making and communication to improve quality of life without increasing costs. The evidence, though weak to moderate in quality, describes mostly positive impacts or no adverse effects. A greater emphasis on patient and caregiver outcomes is needed; and rapid research paradigms, evaluation and implementation studies now need to be prioritised. Future meta-reviews would benefit from looser inclusion criteria to capture other types of reviews containing evidence on emerging innovations such as wearables, smartphone apps, robotics and artificial intelligence.

## Methods

### Search Strategy

The search strategy included the following databases: MEDLINE, MEDLINE In-Process & Other Non-Indexed Citations; EMBASE, PsychINFO, CINAHL, Cochrane Database for Systematic Reviews (CDSR); Cochrane Database of Abstracts of Reviews of Effects (DARE); WHO Global Library (regional indexes only), and Web of Science. The Grey Literature Report (www.greylit.org) was also searched using keywords tailored for this database. The search strategy included MeSH headings and key words related to digital health, palliative care, and technology. All search strategies can be found in the supplementary material online (see Supplementary Material 2). Initial searches were conducted in June 2018; with update searches covering the period from June 2018 to January 2020 conducted in January 2020. Searches were limited to articles published after 2006 to ensure relevance given rapidly evolving technologies. There were no restrictions placed on language.

### Inclusion Criteria

The search strategy targeted systematic reviews explicitly focused on DHIs in palliative care. Systematic reviews of broader healthcare areas which included and separately reported or synthesized studies of DHIs in palliative care were also eligible for inclusion. Using the PICO process,^67^ we defined our target population **(P)** as children and adults who would benefit from palliative care, caregivers (informal and formal), and healthcare professionals delivering palliative care via DHIs or using DHIs to support palliative care decision-making. For the purposes of this review DHIs **(I)**, were defined as approaches in which digital Information and Communication Technologies (ICT) are used to deliver, facilitate or augment palliative care services, including psychological therapies, social support interventions, education, information, anticipatory care planning, remote care support, self-medication/management support, clinical decision support etc. Examples of relevant ICT include telephone, smartphone apps, mobile phones/SMS, videoconferencing, voice over IP (VoIP), instant messaging, email, internet resources, tablets, wearables, electronic patient records. Both synchronous (e.g. videoconferencing) and asynchronous (e.g. email) approaches were included. Our comparator of interest **(C)** was no DHIs or usual care. No limitations were placed on outcomes (**O**), as we were interested in identifying the broad range of outcomes potentially influenced by palliative care DHIs.

### Data extraction

The first and second authors (AF and HO) undertook the database searches and initial screening of titles and abstracts. Where uncertainty existed in relation to potential eligibility, titles and abstracts were independently screened by a third author and ambiguities or disagreements resolved through discussion with the wider team. HO and AF independently assessed papers identified for full-text review, with CP arbitrating where it was unclear whether a review paper should be included. Disagreements and uncertainties were resolved during full team discussions and the authors came to a 100% agreement.

Three co-authors extracted the following information from each of the included systematic reviews: authors, date of publication, country, review aims, search strategy, number of studies included, total number of participants, definition of palliative care, details of participants, functions and medium of DHIs included, reported outcomes, quality assessment methods and conclusions. Three co-authors team then extracted the types of digital health technologies and the intended purposes of the technologies from the individual studies from the included reviews and sought advice from to a fourth co-author in cases of uncertainty.

All reviews were imported into NVivo. Findings from each review were thematically analysed. These themes were then used to structure the results.

### Quality Appraisal

The Assessment of Multiple Systematic Reviews (AMSTAR) checklist was used to critically appraise and score included reviews.^68^ For the purpose of this meta-review the following thresholds were used: low (0-4), moderate (5-8), and high (9-11).^69,70^ Quality appraisal was conducted by four co-authors, with two co-authors independently rating each review. Where disagreements or uncertainties appeared, these were resolved through discussion with the wider team.

### Data Synthesis

We expected substantial heterogeneity amongst systematic reviews as well as amongst the studies included within the reviews. Consequently, we planned to undertake a narrative synthesis. Interpretation was facilitated by discussion amongst the team.

## Data Availability

All data is available on request from the corresponding author.

## Acknowledgements

Thanks to Richard Meade, Marie Curie Head of Public Affairs in Scotland for helpful comments on a draft version of this manuscript. We are very grateful to Marshall Dozier at the University of Edinburgh for advice and assistance running the literature searches.

## Author Contributions

AMF, HOD and CP designed the study. HOD developed the search strategy and conducted the literature searches. HOD, AMF, CS, and JL were involved in data screening and extraction. HOD and AMF synthesized findings and drafted the manuscript. All authors contributed to the final version.

